# Type 1 Diabetes Prevention: a systematic review of studies testing disease-modifying therapies and features linked to treatment response

**DOI:** 10.1101/2023.04.12.23288421

**Authors:** Jamie L. Felton, Kurt J. Griffin, Richard A. Oram, Cate Speake, S. Alice Long, Suna Onengut-Gumuscu, Stephen S. Rich, Gabriela SF Monaco, Carmella Evans-Molina, Linda A. DiMeglio, Heba M. Ismail, Andrea K. Steck, Dana Dabelea, Randi K. Johnson, Marzhan Urazbayeva, Stephen Gitelman, John M. Wentworth, Maria J. Redondo, Emily K. Sims, the ADA/EASD Precision Medicine in Diabetes Initiative Consortium

## Abstract

**Background:** Type 1 diabetes (T1D) results from immune-mediated destruction of insulin-producing beta cells. Efforts to prevent T1D have focused on modulating immune responses and supporting beta cell health; however, heterogeneity in disease progression and responses to therapies have made these efforts difficult to translate to clinical practice, highlighting the need for precision medicine approaches to T1D prevention.

**Methods:** To understand the current state of knowledge regarding precision approaches to T1D prevention, we performed a systematic review of randomized-controlled trials from the past 25 years testing disease-modifying therapies in T1D and/or identifying features linked to treatment response, analyzing bias using a Cochrane-risk-of-bias instrument.

**Results:** We identified 75 manuscripts, 15 describing 11 prevention trials for individuals with increased risk for T1D, and 60 describing treatments aimed at preventing beta cell loss in individuals at disease onset. Seventeen agents tested, mostly immunotherapies, showed benefit compared to placebo (only two prior to T1D onset). Fifty-seven studies employed precision analyses to assess features linked to treatment response. Age, measures of beta cell function and immune phenotypes were most frequently tested. However, analyses were typically not prespecified, with inconsistent methods reporting, and tended to report positive findings.

**Conclusions:** While the quality of prevention and intervention trials was overall high, low quality of precision analyses made it difficult to draw meaningful conclusions that inform clinical practice. Thus, prespecified precision analyses should be incorporated into the design of future studies and reported in full to facilitate precision medicine approaches to T1D prevention.

**Plain Language Summary:** Type 1 diabetes (T1D) results from the destruction of insulin-producing cells in the pancreas, necessitating lifelong insulin dependence. T1D prevention remains an elusive goal, largely due to immense variability in disease progression. Agents tested to date in clinical trials work in a subset of individuals, highlighting the need for precision medicine approaches to prevention. We systematically reviewed clinical trials of disease-modifying therapy in T1D. While age, measures of beta cell function, and immune phenotypes were most commonly identified as factors that influenced treatment response, the overall quality of these studies was low. This review reveals an important need to proactively design clinical trials with well-defined analyses to ensure that results can be interpreted and applied to clinical practice.

## Introduction

Type 1 diabetes (T1D) results from immune-mediated destruction of pancreatic beta cells^1^. Since the discovery of insulin over a century ago, treatment options for persons with type 1 diabetes (T1D) have shown remarkable advancements, including improved insulin formulations, delivery methods, and tools to monitor glycemia ^2^. Even with these transformative advances, considerable negative impacts remain on health outcomes and quality of life ^3–5^. Therapies targeting the underlying pathophysiology of T1D could delay, prevent, or reverse the need for insulin replacement. Many therapies have been proposed and tested as potential agents for disease modification with an ultimate goal of T1D prevention. In 2022, the US Food and Drug Administration approved teplizumab, a monoclonal antibody targeting CD3, as the first therapy to delay the onset of clinical T1D in at-risk individuals ^6^.

Autoimmune beta cell destruction leads to progressive hyperglycemia, abnormal glucose tolerance and eventually clinical T1D diagnosis. This progression is also reflected in C-peptide decline, accelerating in the last preclinical stages before diagnosis and continuing after diagnosis ^7^. Because T1D is an autoimmune disease, most agents tested as potential disease modifying therapies are immunomodulatory, while others target pathologic contributors such as glucose toxicity and beta cell health and function ^8^. Many agents that hold promise in T1D prevention are first tested in individuals at the time of T1D diagnosis because of the more favorable risk-benefit ratio coupled with an increased ease of identifying eligible trial participants.

The Precision Medicine in Diabetes Initiative (PMDI) was established in 2018 by the American Diabetes Association (ADA) in partnership with the European Association for the Study of Diabetes (EASD). The ADA/EASD PMDI includes global thought leaders in precision diabetes medicine who are working to address the burgeoning need for better diabetes prevention and care through precision medicine. This Systematic Review is written on behalf of the ADA/EASD PMDI as part of a comprehensive evidence evaluation of precision prevention in T1D in support of the 2nd International Consensus Report on Precision Diabetes Medicine. The first ADA/ EASD Precision Medicine in Diabetes Consensus Report defined precision prevention as “using information about a person’s unique biology, environment, and/or context to determine their likely responses to health interventions” and states that “precision prevention should optimize the prescription of health-enhancing interventions” ^9^. Given that agents targeting these pathways may have potential adverse effects, and initial therapies may affect efficacy and safety of subsequent treatment approaches, precision medicine is uniquely poised to identify which individuals stand to benefit the most from a given intervention, and to optimize potential risk-benefit ratios for treated-persons. Additionally, once further T1D disease-modifying therapies are approved for clinical use, precision medicine will facilitate selection of therapies guided by the individual’s disease, including potential combination regimens of disease-modifying therapies ^10, 11^.

Therefore, we sought to understand the current state of knowledge regarding precision approaches to T1D disease modification, either to prevent development of clinical disease or its progression. Specifically, we asked if individual characteristics have been robustly identified to select persons for therapeutic optimization of T1D disease-modifying therapies before or at the time of diagnosis. We reviewed and summarized existing trials in this area and identified individual characteristics associated with treatment effects.

## Methods

### Data Source

We developed a search strategy using an iterative process that involved Medical Subject Headings (MeSH) and text words. This search was refined based on a sensitivity check for key articles identified by members of the group (**Supplemental Figure 1**). This strategy was applied to PubMed and EMBASE databases by librarians from Lund University on 2/22/2022.

### Study Selection

The Covidence platform was utilized for stages of systematic review. To qualify for review, studies must have tested a disease-modifying treatment in either initially non-diabetic individuals at risk, or individuals with new onset stage 3 type 1 diabetes (within 1 year of diagnosis). Eligible study types included randomized controlled trials (RCTs); systematic reviews or meta-analyses of RCTs, or post-hoc analyses of RCTs. Selected primary trials or longitudinal follow-up papers of primary trials had a total sample size >=50 and were published as a full paper in English in a peer-reviewed journal within 25 years of the search (2/21/1997-2/22/2022). Papers focusing on a precision approach to identify features associated with a treatment response were also included as long as the total sample size was >10. Longitudinal follow-up papers of RCTs were included if they addressed follow-up data on time to diabetes, C-peptide area under the curve (AUC), or included precision analyses aiming to identify measures or markers of treatment response. Studies were excluded if they included mixed participant populations (i.e. type 1 and type 2 diabetes) or populations with inconsistent definitions across papers (i.e. latent onset diabetes in adults). Several additional key articles identified by the group of experts that also met inclusion criteria but not included in the search results because of search restrictions designed to improve search feasibility were also included in the analysis.

Investigators independently screened and reviewed each potentially relevant article according to preliminary eligibility criteria determined by members of the review team. For Level 1 screening two investigators per article screened each title and abstract. Discordant assessments were discussed and resolved by consensus or arbitration after consultation with a member of the review leadership team (JLF, RO, KJG, MR, or EKS). For Level 2 screening of eligible articles, full texts were retrieved and reviewed by two independent reviewers using the inclusion/exclusion criteria. Discordant assessments were similarly discussed and resolved.

### Data Extraction

Two separate investigators per article extracted data from each article meeting inclusion criteria, with consensus determined by a member of the leadership team. Extracted data included participant characteristics, intervention details, outcomes of intervention on time to diabetes or C-peptide, and methods and findings surrounding precision analyses focused on treatment response. Investigators also performed quality assessments using Covidence’s Cochrane Risk of Bias template in tandem for each eligible study; this included assessments of sequence generation, allocation concealment, masking of participants/personnel, masking of outcome assessment, incomplete outcome data, selective reporting, and any other sources of bias to order to determine overall risk of bias.

### Data Analysis and Synthesis

Because of heterogeneity of clinical interventions (e.g., agent tested, study design, analytical methodology, etc.) we were unable to perform a meta-analysis but instead completed summaries of relevant studies. The protocol of this review was registered at Prospero.com before implementation (available at https://www.crd.york.ac.uk/PROSPEROFILES/310063_PROTOCOL_20221110.pdf).

## Results

### Systematic Review Results

From 1005 studies identified by PubMed and Embase searches, 75 were eligible for extraction (Figure 1). This included original trial papers, trial longitudinal follow-up papers, and papers focused specifically on a precision analysis surrounding treatment response in prevention trials (15 papers from 11 prevention trial cohorts) ^12–26^ and in individuals with new onset T1D (60 total papers from 45 new onset trial cohorts)^27–86^.

**Figure 1.**
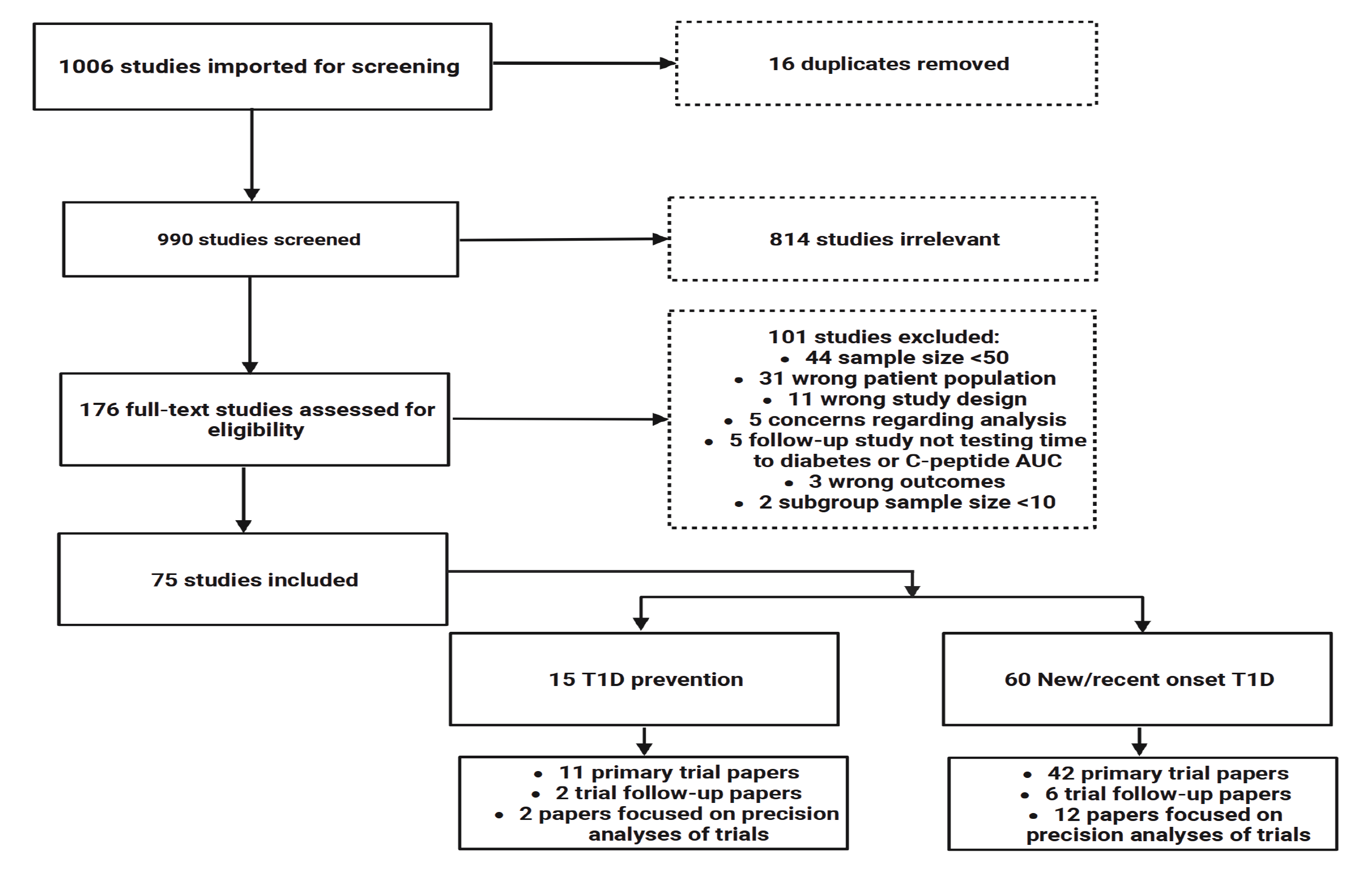
Consort diagram. Flowchart displaying studies screened, and excluded as part of abstract screening, then via full text review/eligibility assessment. 75 total papers were included in extraction. AUC-area under the curve; T1D-Type 1 diabetes

The 15 articles on T1D prevention generated from 11 trials are summarized in **Table 1**. Primary prevention studies in genetically at-risk individuals testing development of islet autoantibodies or time to T1D comprised 27% (3/11) of trials; 63% (7/11) of trials were secondary prevention studies testing effects of interventions in autoantibody positive individuals on time to T1D; one trial tested both genetically at-risk infants and autoantibody positive siblings. Further inclusion criteria for trials included measures of beta cell function, with studies testing antigen-based therapies utilizing specific autoantibody positivity criteria. The DPT-1 oral and parental insulin studies and TrialNet oral insulin study identified participants based on insulin autoantibody positivity and first phase insulin response on intravenous (IV) glucose tolerance testing ^19, 23, 87^. The TrialNet teplizumab prevention study only enrolled individuals with dysglycemia on oral glucose tolerance testing ^88^. Finally, a study testing glutamic acid decarboxylase (GAD) antigen therapy was limited to individuals who were GAD autoantibody positive ^89^. Most prevention trials (9/11; 81%) were multicenter studies; 9/11 (82%) were also double masked, while 2/11 (18%) had no masking. In addition to these 11 papers, two follow-up papers and two papers focused solely on precision analysis of treatment response were also identified (for a total of 15 papers). Overall, only two prevention studies reported a positive impact on time to islet autoantibody positivity or time to diabetes: the primary prevention study testing whey-based hydrolyzed vs cow’s milk formula ^25^ and the secondary prevention study testing teplizumab ^16^ (Fig. 2).

**Figure 2.**
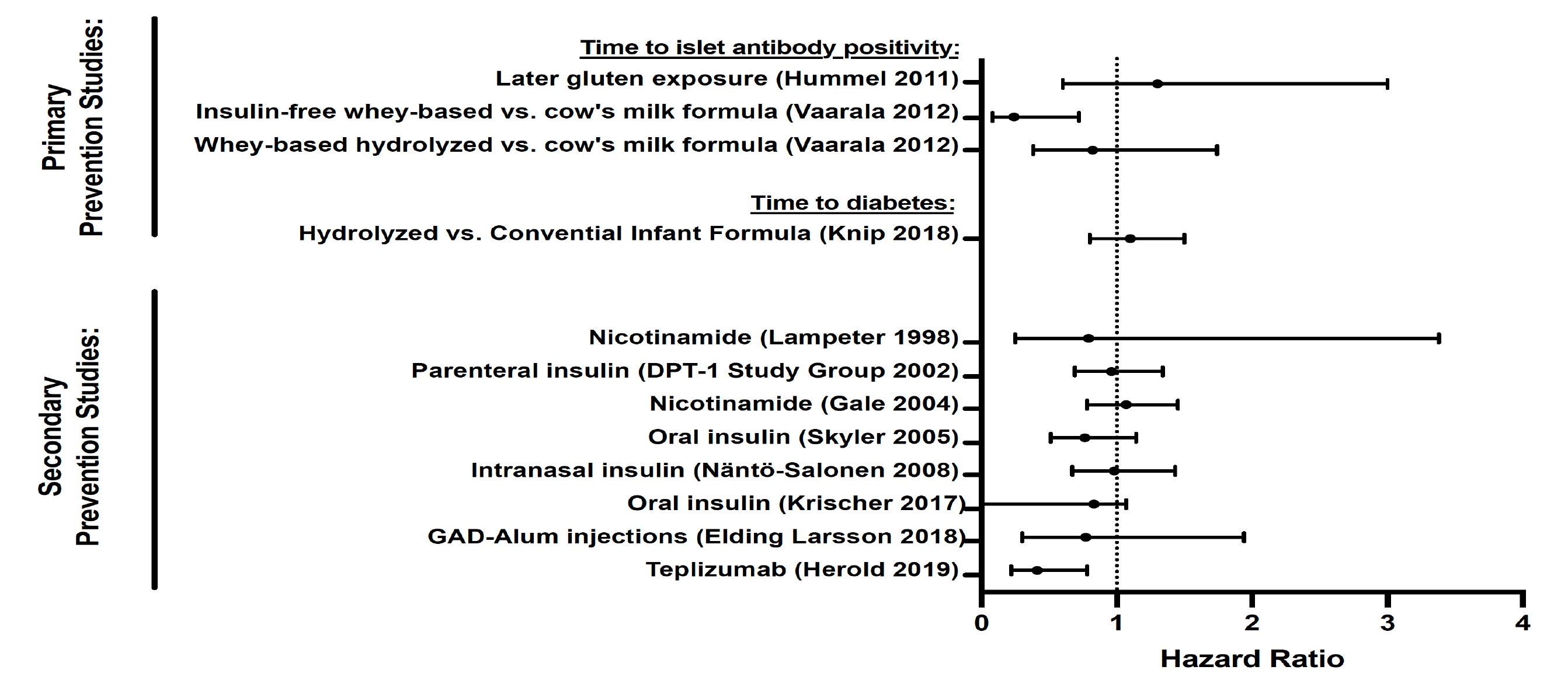
Relative effect of prevention therapies in individuals at-risk for T1D. Forest plot showing hazard ratio for primary prevention studies in genetically at-risk individuals and secondary prevention studies in individuals with elevated islet autoantibody titers. Primary prevention studies are divided by outcome-either time to islet autoantibody positivity or time to diabetes. All secondary prevention studies used time to diabetes as a primary outcome. DPT1 – Diabetes Prevention Trial Type 1; GAD– Glutamic acid decarboxylase

**Table 1.**
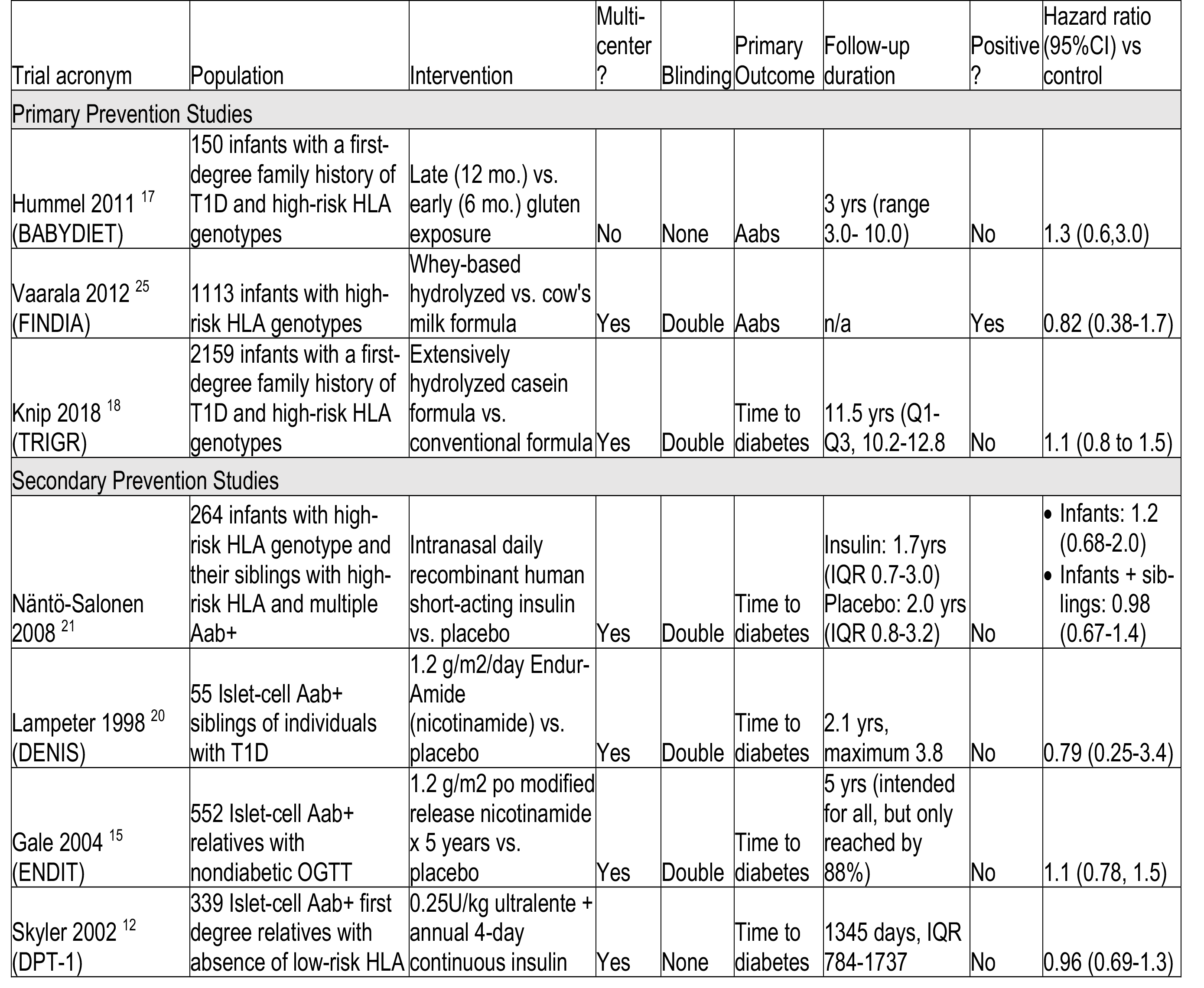
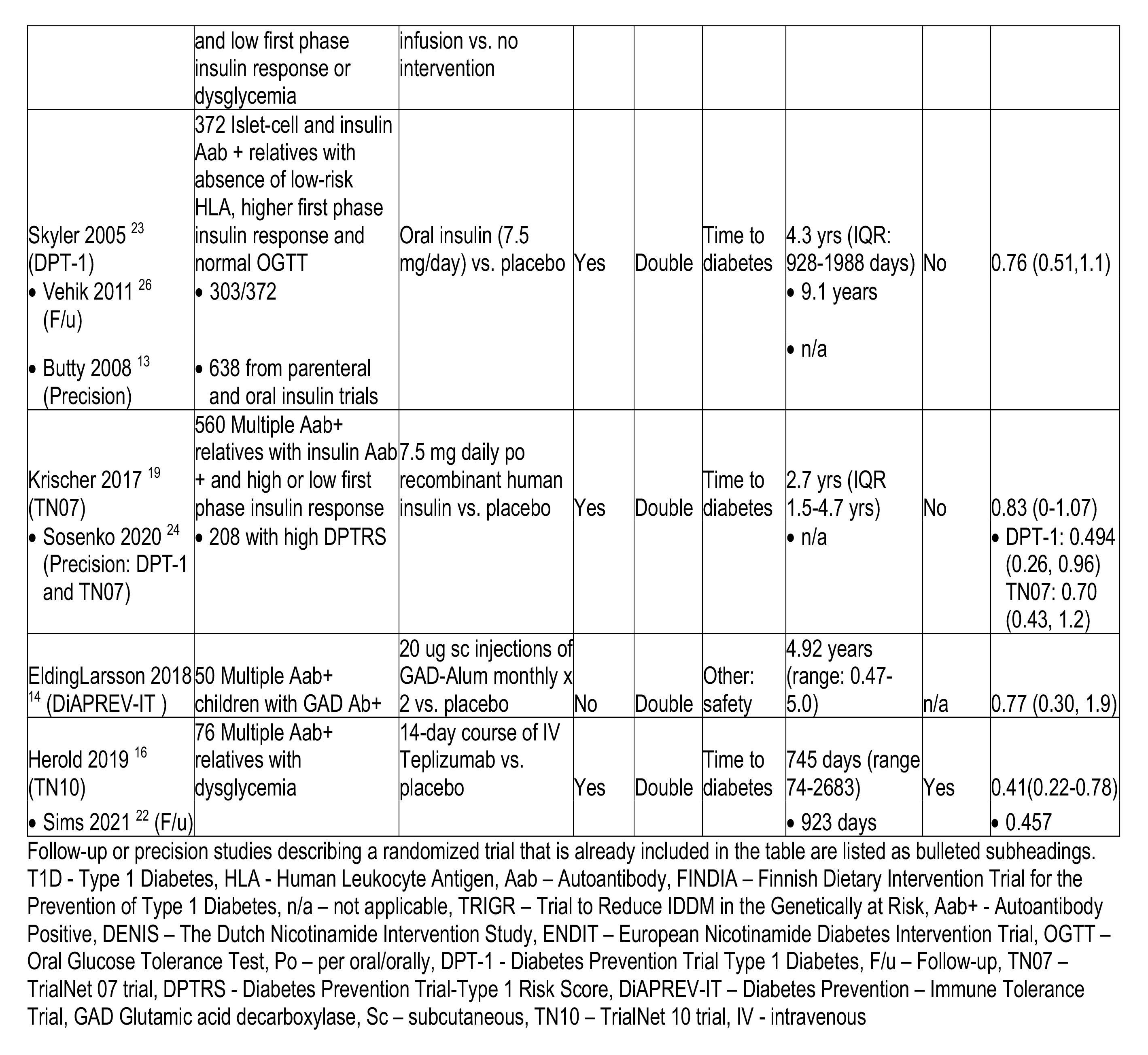
Prevention Studies

The 60 manuscripts generated from 45 trials in the new-onset T1D population included 42 primary trial papers, 6 trial longitudinal follow-up papers and 12 papers focused solely on precision analyses of treatment response (**Fig. 1**). Additional characteristics of these 60 papers are summarized in **Supplemental Table 1**. Here, except for variable age criteria, inclusion criteria were more homogeneous than in prevention studies, typically including participants with a clinical diagnosis of T1D (usually with islet autoantibody positivity) and C-peptide above a certain cutoff. Of the 43 trials, 30 (70%) included both adults and children, 9 (21%) tested only children, and 4 (9%) were performed solely in adults. Five trials had inclusion criteria that included positivity for a specific islet autoantibody. Trials described were typically multicenter studies (39/43; 91%) and double masked (35/43; 81%). Two studies were single masked, two described only masked outcomes testing, three had no masking, and masking was not described in one study.

A measure of beta cell function was by far the most common primary outcome specified amongst new onset trials (single primary outcome in 33/43 (77%), co-primary outcome in 2/43; 5%), although other studies used HbA1c and/or insulin dose and one study used T1D remission. Primary outcome was not specified in 5 trials. All follow-up studies focused on a measure of beta cell function. Trials reporting a measure of beta cell function as the primary outcome most commonly utilized mean C-peptide AUC from a mixed meal tolerance test; values for these data were available for 32/35 primary trials and 5/6 follow up studies and are summarized in **Supplemental Table 2**. Of trial manuscripts reporting these data, less than a fourth identified a positive effect of the intervention on mean C-peptide AUC. These included trials testing imatinib mesylate, low dose anti-thymocyte globulin, teplizumab (anti-CD3 antibody), otelixizumab (anti-CD3 antibody), abatacept (CTLA4-Ig), rituximab (anti-CD20 antibody), golimumab (anti-TNF-alpha), recombinant IFN alpha, and combination of anti-IL-21 antibody with liraglutide.

### Precision analyses focused on features associated with disease-modifying treatment response

To determine whether there were specific features that impacted response to treatment (genetic, metabolic, immune), we assessed papers that included this type of precision analysis. Two papers from prevention and the 12 papers from new onset studies focused solely on precision analyses of treatment response (ie, no analysis of primary trial or longitudinal follow-up analysis of primary trial). An additional 43 papers also included some aspect of precision analysis (**summarized in Supplemental Table 3**). As shown in **Fig. 3A**, of 57 total papers identified, most (38/57; 67%) were primary trial papers with a section focused on features of treatment response. Just over half of the primary trial follow-up papers (5/8; 63%) included precision analyses of treatment response.

**Figure 3.**
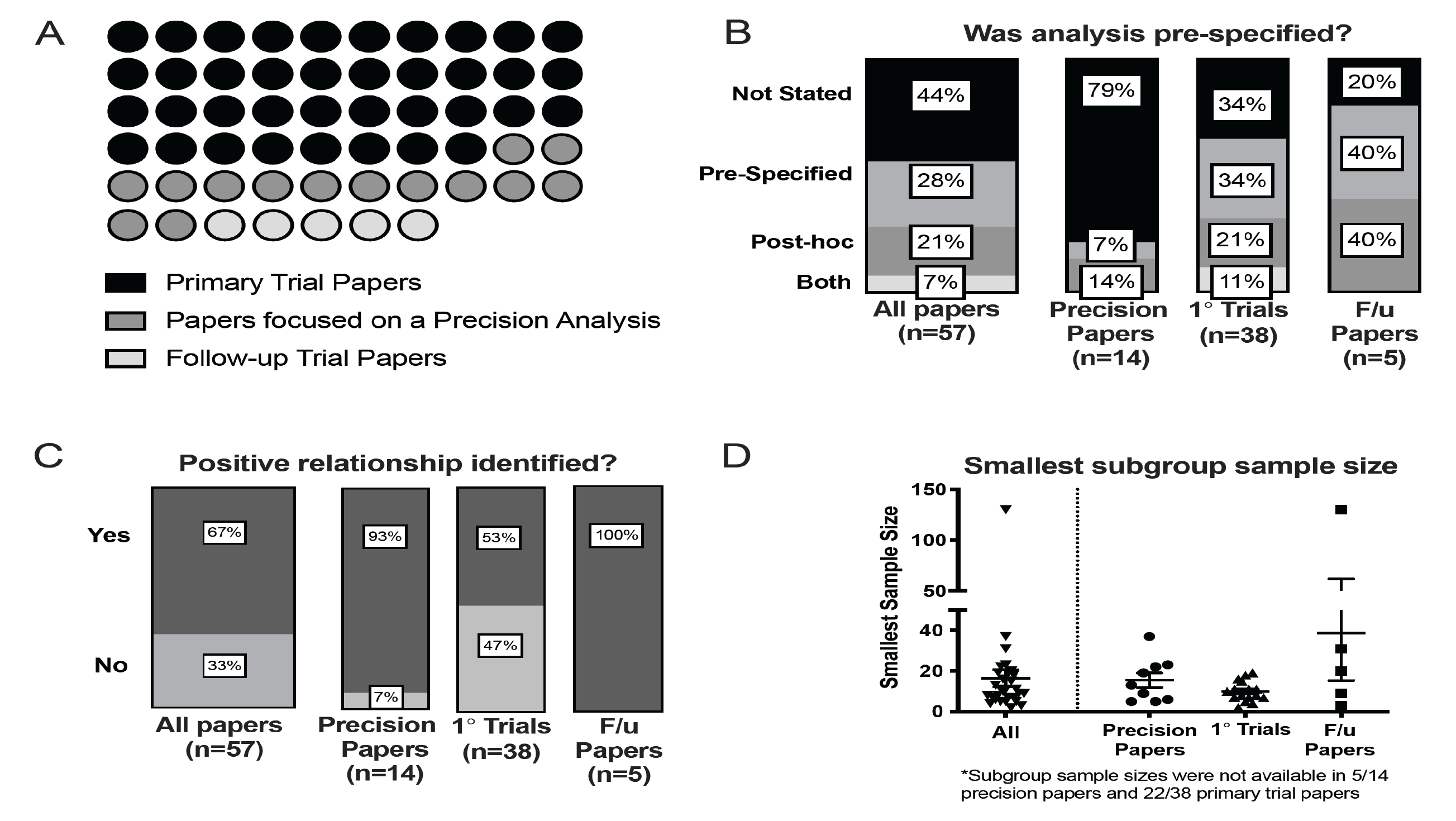
Precision analyses focused on treatment response were mostly part of primary trial papers, tended to be post-hoc, and were biased toward positive findings. **A.** Dot plot showing breakdown of the 57 papers that included precision analyses. These were 67% primary trial papers, 24% manuscripts that focused on precision analyses related to treatment response, and 9% longitudinal follow-up papers presenting updates on primary trial results. **B.** Stacked bar graphs showing relative frequencies of papers with precision analyses that were defined as prespecified in the manuscript text. **C.** Stacked bar graph displaying relative frequencies of papers reporting positive findings related to associations with treatment effects. **D.** For papers that listed sample sizes of subgroups tested for differential treatment effects (only 53% of all papers with precision analyses), the smallest samples size reported is displayed, with mean and SEM indicated. F/u-follow-up

While precision analysis of treatment response was commonly reported, this was rarely pre-specified, occurring in just 16/57 (28**%**) of papers studied (**Fig. 3B**). Prespecified precision analyses were more common in primary trial or primary trial follow-up papers. For primary trials, 34% (13/38) of precision analyses were prespecified, and 10.5% (4/38) had both pre-specified and post-hoc analyses. For follow-up papers 40% (2/5) were pre-specified. In contrast, only 7% (1/14) of papers focused specifically on precision analyses described a prespecified analysis plan. Analyses tended to identify a positive relationship with treatment effect (**Fig. 3C**), with 37/57 (67%) studies identifying a significant relationship between a feature and treatment response. This was more prevalent for precision analyses in primary trial follow-up papers (5/5; 100%) and in precision analysis-only papers (13/14; 93%).

Because sample sizes inevitably decrease as groups are subdivided for precision analyses, we next looked at sample sizes for the precision subgroups. Only slightly over half (30/57) of papers reported sample sizes for all subgroups defined by precision features. Within these 30 manuscripts, we observed wide variability in sample sizes of subgroups analyzed. **Fig. 3D** displays reported values for the smallest subgroup sample size described. Overall median values were 11 (interquartile range of 7-19) participants, and ranged from 2-128 participants.

**Fig. 4A** displays the number of precision features tested for each paper. For all papers, the median number of features tested was 3 (interquartile range of 1-7). This tended to be higher in papers focused solely on precision analyses (median of 6.5 with several papers testing numerous subgroups as part of sequencing, array, or flow cytometry analysis). Forty-one papers analyzed multiple precision features. Of these applicable analyses, corrections for multiple comparisons were either not mentioned or not performed in 35/41 (85%) of papers, particularly for trials (100% of applicable papers with multiple comparisons not described or not performed) (**Figure 4B**).

**Figure 4.**
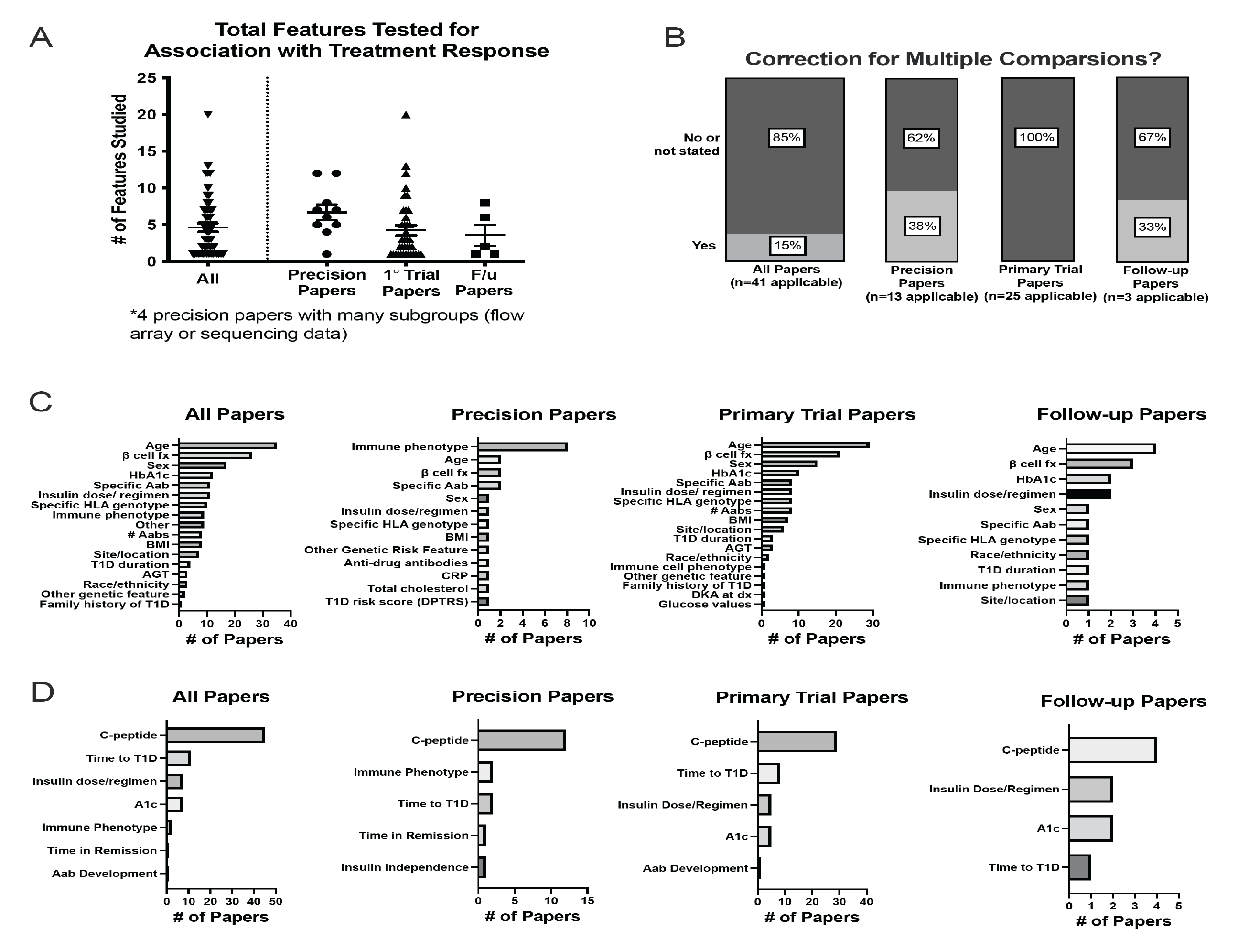
Precision analyses tested many features, most commonly age and beta cell function, infrequently corrected for multiple comparisons, and typically tested for differential impacts on a C-peptide based measure. **A.** Total number of features tested for association with each treatment response, with mean and SEM indicated. **B.** Stacked bar graph showing relative frequencies of papers that did or did not correct for multiple comparisons. **C.** Relative frequencies of individual features tested for associations with treatment response. **D.** Relative frequencies of outcomes utilized to assess for the presence of any features associated with differential treatment response. F/u – follow-up, fx – function, Hba1c – Hemoglobin A1c, Aab – Autoantibody, HLA – Human Leukocyte Antigen, BMI – Body Mass Index, T1D – Type 1 Diabetes, AGT – Abnormal Glucose Tolerance, CRP – C-reactive Protein, DPTRS – Diabetes Prevention Trial-Type 1 Risk Score, DKA – Diabetes Ketoacidosis, Dx - diagnosis

We next examined the types of features tested for relationships with treatment response (**Fig. 4C**). In trial papers and follow up papers, age was most commonly tested (>3/4 of analyses), followed by a measure of beta cell function (>1/2 of analyses). Only 9/36 (25%) studies testing age identified a significant relationship with treatment response; these were all in the new onset period ^27, 36, 41, 43, 49, 53, 56, 79, 82^. Here, younger age groups showed improved treatment responses to teplizumab, ChAglyCD3, and Vitamin E ^36, 49, 53, 56, 79, 82^, In contrast older age was linked to a beneficial treatment response vs. placebo with high-dose antithymocyte globulin (ATG) and oral insulin (both studies with negative findings overall) ^41, 43^. One study showed that younger age was linked to a more rapid decline of C-peptide compared to placebo in Bacillus Calmette-Guerin (BCG) vaccine-treated individuals ^27^. Baseline measures of beta cell function were linked to differences in treatment response in 10/26 (38%) of analyses where this relationship was tested ^16, 19, 35, 42, 49, 55, 56, 68, 83, 84^. In two papers focused on prevention studies, measures linked to worsened beta cell function were associated with an improved response to treatment (with oral insulin or teplizumab)^16, 19^. Analyses testing trials in the new-onset period had split results: teplizumab, ChAglyCD3, linomide, and atorvastatin performed better compared to placebo in groups with better baseline beta cell function measures ^35, 49, 55, 56, 83^. In contrast, canakinumab, imatinib mesylate, and the anti-IL-21/liraglutide combination showed stronger treatment effects in individuals with lower baseline beta cell function measures ^42, 68, 84^. Taken in aggregate these results highlight evidence that baseline beta cell function may impact treatment response, but the direction of impact likely varies by treatment used and stage of disease.

Interestingly, in contrast to primary trial papers, precision papers most commonly tested relationships of an immune cell phenotype with treatment response (57%). Because only two papers identified included a favorable response to time to type 1 diabetes diagnosis, treatment response was assessed using a range of alternative outcomes (**Fig. 4D**). For all types of papers, a measure of C-peptide was most commonly used as an alternative outcome to gauge treatment response (range of 44-68%).

### Risk of Bias/Quality Assessments

A finding impacting studies in all categories was a lack of racial and ethnic diversity in participant populations. Data on participant race was available in less than a third (23/75) of total papers; for reported papers, participants self-reporting as white race comprised a median of 92% of the total study population (interquartile range of 88- 96%). Ethnicity was reported in 20 papers; within these manuscripts, participants self-reporting as identifying with a Hispanic ethnicity comprised a median of 5% of study participants (interquartile range of 3-9%).

When assessing additional risks of bias, we found that many papers did not include details sufficient to assess these risks (**Fig. 5**). Although over half of primary trial papers were considered to utilize high quality methods for sequence generation and allocation concealment, 32-37% did not describe methods adequately for assessment. Follow-up and precision papers infrequently described these methods, commonly citing a primary trial paper instead (75-100%). Blinding was described more frequently, with at least double blinding in 63-74% of follow-up and primary trial papers, although 23-25% had single or no blinding. In contrast, blinding of outcome assessments was either not described or did not occur in 79% of primary trial papers. Most precision papers referenced primary papers and so blinding was challenging to assess. Completeness of outcome data reporting was assessed by considering reasons and numbers for attrition or exclusion in studies. Reporting of outcome was overall high quality for trials and follow-up studies (75-79%). This was less frequently the case for precision papers, only half of which reported on reasons for incomplete outcome data. While the large majority (87%) of trial papers described a prespecified primary endpoint, only 75% of follow-up papers and 21% of precision papers solely included analyses that were noted to be prespecified. Additional sources of bias were identified in 33/75 total papers (44%), these biases were also acknowledged by study authors. These were most frequently acknowledged funding or support by a pharmaceutical company. However, another source of bias that was not addressed as a limitation by the authors was identified in 3 papers (all primary trial papers). No concerns for other unacknowledged sources of bias were identified in follow-up studies and precision studies.

**Figure 5.**
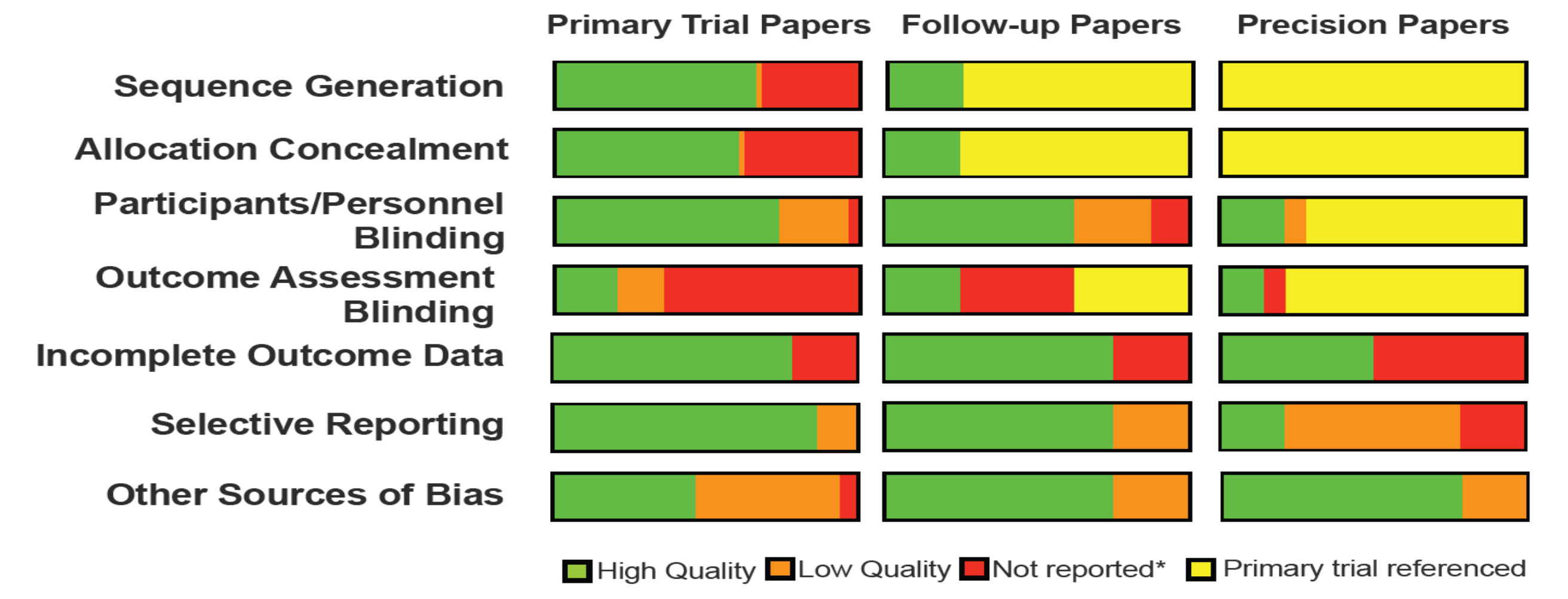
Risk of bias assessments for each paper category. Bias was assessed using Covidence’s Cochrane risk of bias tool. For sequence generation, allocation concealment, and blinding categories, raters had the option of selecting high quality (green), low quality (orange), not reported (red), or that a decision could not be made because of primary trial was referenced in methods (yellow). For incomplete outcome data, raters only had the option to choose high quality/ data provided (green) or low quality/ data not provided (red). For selective reporting, raters had the option to select high quality/primary endpoint predefined (green), low quality/primary endpoint not defined (orange) or low quality/not reported (red). For other sources of bias, raters had the option to select high quality/none (green), low quality/bias present but identified and considered (orange), or low quality/obvious bias present and not addressed (red).

## Conclusions

We systematically reviewed 25 years of large (n>=50 participants) randomized controlled trials focused on disease modifying agents in T1D and analyses focused on identifying features of treatment response to these agents. Our search identified 17 agents, mostly immunotherapies, that have shown benefit compared to placebo (though only two prior to clinical diabetes onset). We found most precision subgroup analyses were included as components of manuscripts reporting primary outcomes of trials. The analyses most commonly focused on age or a measure of beta cell function as potential features that may identify individuals with better responses to specific disease-modifying therapies. We also identified stand-alone papers focused on understanding treatment response. Immune phenotypes were most commonly assessed as features to define heterogeneity of treatment response in these papers.

Time to T1D was the most consistent primary outcome of T1D prevention studies, but inclusion criteria for these studies varied widely across trials and included genetic risk, presence of islet autoantibodies, and changes in glycemia and/or beta cell function. Most intervention studies were in new-onset T1D and the vast majority of these new-onset studies analyzed impacts on C-peptide AUC during a mixed meal test as a primary outcome, consistent with the consensus recommendations by Palmer and colleagues ^90^. Most new-onset studies identified had similar inclusion criteria, with ∼10% of studies specifying a primary analysis of a more precise subgroup of individuals for study based on specific autoantibody positivity. The majority of trials at onset of disease likely reflects the longstanding precedent of defining T1D by established clinical criteria (established due to the link with future risk of complications)^91^. A key consideration is the appealing risk-benefit balance of giving immunotherapy to individuals with a clinical T1D diagnosis, compared to persons at-risk who may never go on to develop T1D. Recent progress in understanding the natural history of T1D, particularly the high risk associated with progression from multiple autoantibodies to clinical T1D ^92^, has led to a definition of stage 1 and stage 2 defined by the presence of autoantibodies and dysglycemia respectively ^93, 94^. The combination of defining pre-clinical stages 1 and 2 and the recent positive trial of teplizumab in stage 2 may change the balance of prevention versus new onset trials in the future.

A challenge within the field has been a clear and consistent delineation of the definition of intervention “responders” within larger trial populations. Strategies have included consideration of time to diabetes, insulin use, stratification based on changes in C-peptide, and identification of individuals exhibiting less loss of C-peptide based on placebo controls ^16, 24, 56, 62^. Our review suggests that C-peptide is by far the most frequent outcome measure used to identify differential treatment responses, but approaches to stratify based on C-peptide were highly variable. Consistent approaches, such as a quantifiable metric based on expected values ^95^ will allow better comparison of features associated with treatment response across trials.

Age and measures of beta cell function were most frequently identified as factors associated with differential treatment response in primary trial and primary trial follow-up papers. For example, younger age was linked to an improved treatment response in several new-onset trials using CD3-based agents ^49, 53, 56, 82^. The association of age with treatment response is in keeping with the strong associations of age to features of T1D in many observational and natural history studies, before and after clinical diagnosis ^10, 96–98^. Differences in pancreas histology have been identified in donors with younger age of diagnosis ^99, 100^. However, it is unclear whether differences in treatment response linked to age are associated with differences in underlying disease pathophysiology vs. differences in severity or progression of disease at the time of treatment. The observation that age differentially impacts outcomes in different trials, in addition to stratification of both immune phenotypes and beta cell function by age, supports the idea that the underlying biological reasons for age associations could be linked to mechanism and are important to consider in future trial designs and potentially, in future precision therapy.

Importantly, 38% of studies testing impacts of baseline beta cell function showed a significant link to treatment response, consistent with the substantial body of literature identifying an ongoing dialogue between autoimmunity and the beta cell in T1D ^96, 101–108^. Interestingly, findings somewhat differed depending on stage of intervention. Here, two unique prevention studies testing oral insulin and teplizumab showed that worse beta cell function was associated with improved treatment outcomes compared to placebo ^16, 19^. In contrast, CD3-based therapy trials after disease onset showed an association between higher measures of beta cell function and improved outcomes ^49, 55, 56^. These differences highlight the importance of considering disease stage in design and interpretation of intervention efforts ^109^. Especially at earlier stages in the disease process, abnormalities in beta cell function could allow insight into a therapeutic window during active disease or immune attack, and optimal timing of therapy ^110^. In contrast, in more advanced disease after diagnosis, associations with differences in beta cell function could reflect differences in the degree of disease progression, and so amenability to prolonged preservation of a larger residual beta cell mass. Differences in the relationships between beta cell function measures and outcomes for different agents in the new onset period also highlight agent mechanism of action as a critical consideration for designs incorporating beta cell function into stratification of trial populations and precision approaches to disease-modifying therapy.

Specific autoantibodies and immune cell phenotypes were also linked to treatment response for multiple agents. An important consideration in these types of assays is reproducibility. Harmonization to international criteria would facilitate cross-study comparisons and improve reproducibility, a critical challenge with functional immune markers. The T1D field has been strengthened by an international standardization program for autoantibody measurement that underpinned the development of type 1 diabetes staging criteria ^111^. An important consideration is that if novel mechanistic markers (immune, metabolic, or other) can be used to predict treatment response, then similar scrutiny and standardization of these markers will be needed. Pragmatic approaches to biomarker development need to include considerations of reproducibility to be successfully implemented.

Most studies reviewed did not report data on race or ethnicity; for those that did report these data, populations studied largely identified as non-Hispanic white. Barriers in screening of traditionally underrepresented populations is a recognized issue amongst T1D natural history and intervention studies ^95, 112^. This is especially important to address moving forward given the rising incidence of T1D in these populations ^113^.

Our analysis identified important methodologic considerations with many precision analyses. Most trial manuscripts (primary or follow-up) included precision analyses that were not prespecified. Corrections for multiple comparisons were rare. Additionally, subgroup sizes were infrequently reported, but when available, these group sizes were highly variable and as small as n=2 participants. Papers also tended to show positive results, raising concern for publication bias.

While these issues are a known limitation of hypothesis-generating exploratory analyses, follow-up studies focusing on testing these positive relationships a priori will be critical to the application of clinically meaningful precision medicine. An example of the necessity of hypothesis testing was the TrialNet oral insulin prevention study, which was prospectively designed to test a responder subgroup identified in the Diabetes Preventional Trial Type 1 (DPT-1) trial with high insulin autoantibody titers, and ultimately found no significant impact of treatment within this group ^114^. Another example of a trial moving forward with prospective testing based on subgroup analyses is the DIAGNODE 3 study, which will prospectively test intralymphatic GAD-alum injections in the HLA DR3-DQ2 population (NCT05018585). This approach is based on a meta-analysis of subcutaneous GAD-alum trials ^115^ and a small study of intralymphatic injections showing a preferential benefit vs. placebo in this population ^66^. These studies were not included in the current review due to participant time from diagnosis and sample size. Prospective testing of potential responder subgroups is needed to validate findings before they can be integrated into precision approaches. Trials designed to limit participant heterogeneity based on features associated with treatment response may ultimately allow for clearer determinations of effect, and a greater number of positive trials.

This study had limitations. For feasibility, we restricted our review of primary trials to those enrolling a minimum of 50 total participants. Because of this, some trials were not reviewed, including positive trials testing alefacept ^116, 117^ and verapamil in the new onset period ^118^. A large pediatric follow-up trial testing verapamil (positive outcome) and tight metabolic control with hybrid closed loop (negative outcome) was published after conclusion of our systematic review ^119, 120^.

In summary, our review identified significant progress towards defining effective disease-modifying therapies for T1D. Overall this work highlights the impact of consensus agreement on trial outcomes to allow between trial outcomes comparisons and standardization of precision measures to study subgroups of patients or at-risk individuals. Although many associations of interest have been identified, the impact and clinical utility of these observations is weakened by post-hoc study design. Pre-specified adequately powered subgroup analyses focused on age, beta cell function, HLA genotypes, and immune measures will allow stronger conclusions from future studies and should be considered when planning trials. Finally, reports of future trials would benefit from including adequate details to assess potential risks of bias.

## Data Availability

All studies reviewed were identified via publically available databases (PubMed and Embase). Article review data supporting the findings of this study are available upon reasonable request from the corresponding author.

## Acknowledgments

We thank Krister Aronsson and Maria Bjorklund from Lund University for assistance with database searches and Russell de Souza from McMaster University for advice on critical appraisal.

## Author contributions

JLF, KJG, RAO, MJR and EKS designed the project, performed systematic review, interpreted results, and wrote and edited the manuscript. CS, SAL, SOG, SSR, GSFM, CEM, LAD, HMI, AKS, DD, RKJ, MU, SG, and JMW contributed to design of the project, performed systematic review, and edited the manuscript.

## Competing Interests

EKS has received compensation for educational lectures from Medscape, ADA, and MJH Life Sciences and as a consultant for DRI Healthcare. CEM reported serving on advisory boards for Provention Bio, Isla Technologies, MaiCell Technologies, Avotres, DiogenyX, and Neurodon; receiving in-kind research support from Bristol Myers Squibb and Nimbus Pharmaceuticals; and receiving investigator-initiated grants from Lilly Pharmaceuticals and Astellas Pharmaceuticals. LAD reports research support to institution from Dompe, Lilly, Mannkind, Provention, Zealand and consulting relationships with Abata and Vertex. RAO had a UK MRC Confidence in concept grant to develop a T1D GRS biochip with Randox Ltd, and has ongoing research funding from Randox R & D. No other authors report any relevant conflicts of interest.

Data Availability statement all studies reviewed were identified via publically available databases (PubMed and Embase). Article review data supporting the findings of this study are available upon reasonable request from the corresponding author.

## References

1 DiMeglio, L. A., Evans-Molina, C. & Oram, R. A. Type 1 diabetes. Lancet 391, 2449–2462 (2018). https://doi.org:10.1016/S0140-6736(18)31320-5

2 Sims, E. K., Carr, A. L. J., Oram, R. A., DiMeglio, L. A. & Evans-Molina, C. 100 years of insulin: celebrating the past, present and future of diabetes therapy. Nat Med 27, 1154–1164 (2021). https://doi.org:10.1038/s41591-021-01418-2

3 Livingstone, S. J. et al. Estimated life expectancy in a Scottish cohort with type 1 diabetes, 2008-2010. JAMA 313, 37-44 (2015). https://doi.org:10.1001/jama.2014.16425

4 Pambianco, G. et al. The 30-year natural history of type 1 diabetes complications: the Pittsburgh Epidemiology of Diabetes Complications Study experience. Diabetes 55, 1463–1469 (2006). https://doi.org:10.2337/db05-1423

5 Sussman, M., Benner, J., Haller, M. J., Rewers, M. & Griffiths, R. Estimated Lifetime Economic Burden of Type 1 Diabetes. Diabetes Technol Ther 22, 121–130 (2020). https://doi.org:10.1089/dia.2019.0398

7. FDA Approves First Drug That Can Delay Onset of Type 1 Diabetes, <https://www.fda.gov/news-events/press-announcements/fda-approves-first-drug-can-delay-onset-type-1-diabetes> (

7 Bogun, M. M., Bundy, B. N., Goland, R. S. & Greenbaum, C. J. C-Peptide Levels in Subjects Followed Longitudinally Before and After Type 1 Diabetes Diagnosis in TrialNet. Diabetes Care 43, 1836–1842 (2020). https://doi.org:10.2337/dc19-2288

8 Besser, R. E. J. et al. ISPAD clinical practice consensus guidelines 2022: Stages of type 1 diabetes in children and adolescents. Pediatr Diabetes (2022). https://doi.org:10.1111/pedi.13410

9 Chung, W. K. et al. Precision medicine in diabetes: a Consensus Report from the American Diabetes Association (ADA) and the European Association for the Study of Diabetes (EASD). Diabetologia 63, 1671–1693 (2020). https://doi.org:10.1007/s00125-020-05181-w

10 Battaglia, M. et al. Introducing the Endotype Concept to Address the Challenge of Disease Heterogeneity in Type 1 Diabetes. Diabetes Care 43, 5–12 (2020). https://doi.org:10.2337/dc19-0880

11 Anderson, R. L. et al. Innovative Designs and Logistical Considerations for Expedited Clinical Development of Combination Disease-Modifying Treatments for Type 1 Diabetes. Diabetes Care 45, 2189–2201 (2022). https://doi.org:10.2337/dc22-0308

13. Effects of insulin in relatives of patients with type 1 diabetes mellitus. N Engl J Med 346, 1685-1691 (2002). https://doi.org:10.1056/NEJMoa012350

13 Butty, V., Campbell, C., Mathis, D. & Benoist, C. Impact of diabetes susceptibility loci on progression from pre-diabetes to diabetes in at-risk individuals of the diabetes prevention trial-type 1 (DPT-1). Diabetes 57, 2348–2359 (2008). https://doi.org:10.2337/db07-1736

15. Elding Larsson, H., Lundgren, M., Jonsdottir, B., Cuthbertson, D. & Krischer, J. Safety and efficacy of autoantigen-specific therapy with 2 doses of alum-formulated glutamate decarboxylase in children with multiple islet autoantibodies and risk for type 1 diabetes: A randomized clinical trial. Pediatr Diabetes 19, 410–419 (2018). https://doi.org:10.1111/pedi.12611

15 Gale, E. A., Bingley, P. J., Emmett, C. L. & Collier, T. European Nicotinamide Diabetes Intervention Trial (ENDIT): a randomised controlled trial of intervention before the onset of type 1 diabetes. Lancet 363, 925–931 (2004). https://doi.org:10.1016/s0140-6736(04)15786-3

16 Herold, K. C. et al. An Anti-CD3 Antibody, Teplizumab, in Relatives at Risk for Type 1 Diabetes. New England Journal of Medicine 381, 603-613 (2019). https://doi.org:10.1056/nejmoa1902226

17 Hummel, S., Pflüger, M., Hummel, M., Bonifacio, E. & Ziegler, A. G. Primary dietary intervention study to reduce the risk of islet autoimmunity in children at increased risk for type 1 diabetes: the BABYDIET study. Diabetes Care 34, 1301–1305 (2011). https://doi.org:10.2337/dc10-2456

18 Knip, M. et al. Effect of Hydrolyzed Infant Formula vs Conventional Formula on Risk of Type 1 Diabetes. JAMA 319, 38 (2018). https://doi.org:10.1001/jama.2017.19826

19 Krischer, J. P., Schatz, D. A., Bundy, B., Skyler, J. S. & Greenbaum, C. J. Effect of Oral Insulin on Prevention of Diabetes in Relatives of Patients With Type 1 Diabetes: A Randomized Clinical Trial. Jama 318, 1891–1902 (2017). https://doi.org:10.1001/jama.2017.17070

20 Lampeter, E. F. et al. The Deutsche Nicotinamide Intervention Study: an attempt to prevent type 1 diabetes. DENIS Group. Diabetes 47, 980–984 (1998). https://doi.org:10.2337/diabetes.47.6.980

21 Näntö-Salonen, K. et al. Nasal insulin to prevent type 1 diabetes in children with HLA genotypes and autoantibodies conferring increased risk of disease: a double-blind, randomised controlled trial. Lancet 372, 1746–1755 (2008). https://doi.org:10.1016/s0140-6736(08)61309-4

22 Sims, E. K. et al. Teplizumab improves and stabilizes beta cell function in antibody-positive high-risk individuals. Sci Transl Med 13 (2021). https://doi.org:10.1126/scitranslmed.abc8980

23 Skyler, J. S. et al. Effects of oral insulin in relatives of patients with type 1 diabetes: The Diabetes Prevention Trial--Type 1. Diabetes Care 28, 1068–1076 (2005). https://doi.org:10.2337/diacare.28.5.1068

24 Sosenko, J. M. et al. Slowed Metabolic Decline After 1 Year of Oral Insulin Treatment Among Individuals at High Risk for Type 1 Diabetes in the Diabetes Prevention Trial-Type 1 (DPT-1) and TrialNet Oral Insulin Prevention Trials. Diabetes 69, 1827–1832 (2020). https://doi.org:10.2337/db20-0166

25 Vaarala, O. et al. Removal of bovine insulin from cow’s milk formula and early initiation of beta-cell autoimmunity in the FINDIA pilot study. Archives of Pediatrics and Adolescent Medicine 166, 608–614 (2012). https://doi.org:10.1001/archpediatrics.2011.1559

26 Vehik, K. et al. Long-term outcome of individuals treated with oral insulin: diabetes prevention trial-type 1 (DPT-1) oral insulin trial. Diabetes Care 34, 1585–1590 (2011). https://doi.org:10.2337/dc11-0523

27 Allen, H. F. et al. Effect of Bacillus Calmette-Guerin vaccination on new-onset type 1 diabetes. A randomized clinical study. Diabetes Care 22, 1703–1707 (1999). https://doi.org:10.2337/diacare.22.10.1703

28 Ambery, P. et al. Efficacy and safety of low-dose otelixizumab anti-CD3 monoclonal antibody in preserving C-peptide secretion in adolescent type 1 diabetes: DEFEND-2, a randomized, placebo-controlled, double-blind, multi-centre study. Diabet Med 31, 399–402 (2014). https://doi.org:10.1111/dme.12361

29 Aronson, R. et al. Low-dose otelixizumab anti-CD3 monoclonal antibody DEFEND-1 study: results of the randomized phase III study in recent-onset human type 1 diabetes. Diabetes Care 37, 2746–2754 (2014). https://doi.org:10.2337/dc13-0327

30 Ataie-Jafari, A. et al. A randomized placebo-controlled trial of alphacalcidol on the preservation of beta cell function in children with recent onset type 1 diabetes. Clin Nutr 32, 911–917 (2013). https://doi.org:10.1016/j.clnu.2013.01.012

31 Buckingham, B. et al. Effectiveness of early intensive therapy on β-cell preservation in type 1 diabetes. Diabetes Care 36, 4030–4035 (2013). https://doi.org:10.2337/dc13-1074

32 Cabrera, S. M. et al. Innate immune activity as a predictor of persistent insulin secretion and association with responsiveness to CTLA4-Ig treatment in recent-onset type 1 diabetes. Diabetologia 61, 2356–2370 (2018). https://doi.org:10.1007/s00125-018-4708-x

33 Chaillous, L. et al. Oral insulin administration and residual beta-cell function in recent-onset type 1 diabetes: a multicentre randomised controlled trial. Diabète Insuline Orale group. Lancet 356, 545–549 (2000). https://doi.org:10.1016/s0140-6736(00)02579-4

34 Christie, M. R., Mølvig, J., Hawkes, C. J., Carstensen, B. & Mandrup-Poulsen, T. IA-2 antibody-negative status predicts remission and recovery of C-peptide levels in type 1 diabetic patients treated with cyclosporin. Diabetes Care 25, 1192–1197 (2002). https://doi.org:10.2337/diacare.25.7.1192

35 Coutant, R. et al. Low dose linomide in Type I juvenile diabetes of recent onset: a randomised placebo-controlled double blind trial. Diabetologia 41, 1040–1046 (1998). https://doi.org:10.1007/s001250051028

36 Crinò, A. et al. A randomized trial of nicotinamide and vitamin E in children with recent onset type 1 diabetes (IMDIAB IX). Eur J Endocrinol 150, 719–724 (2004). https://doi.org:10.1530/eje.0.1500719

37 Demeester, S. et al. Preexisting insulin autoantibodies predict efficacy of otelixizumab in preserving residual β-cell function in recent-onset type 1 diabetes. Diabetes Care 38, 644–651 (2015). https://doi.org:10.2337/dc14-1575

38 Diggins, K. E. et al. Exhausted-like CD8+ T cell phenotypes linked to C-peptide preservation in alefacept-treated T1D subjects. JCI Insight 6 (2021). https://doi.org:10.1172/jci.insight.142680

39 Eichmann, M. et al. Costimulation Blockade Disrupts CD4(+) T Cell Memory Pathways and Uncouples Their Link to Decline in beta-Cell Function in Type 1 Diabetes. J Immunol 204, 3129–3138 (2020). https://doi.org:10.4049/jimmunol.1901439

40 Enander, R. et al. Beta cell function after intensive subcutaneous insulin therapy or intravenous insulin infusion at onset of type 1 diabetes in children without ketoacidosis. Pediatric Diabetes 19, 1079–1085 (2018). https://doi.org:10.1111/pedi.12657

41 Ergun-Longmire, B. et al. Oral insulin therapy to prevent progression of immune-mediated (type 1) diabetes. Ann N Y Acad Sci 1029, 260–277 (2004). https://doi.org:10.1196/annals.1309.057

42 Gitelman, S. E. et al. Imatinib therapy for patients with recent-onset type 1 diabetes: a multicentre, randomised, double-blind, placebo-controlled, phase 2 trial. Lancet Diabetes Endocrinol 9, 502–514 (2021). https://doi.org:10.1016/s2213-8587(21)00139-x

43 Gitelman, S. E. et al. Antithymocyte globulin therapy for patients with recent-onset type 1 diabetes: 2 year results of a randomised trial. Diabetologia 59, 1153–1161 (2016). https://doi.org:10.1007/s00125-016-3917-4

44 Gitelman, S. E. et al. Antithymocyte globulin treatment for patients with recent-onset type 1 diabetes: 12-month results of a randomised, placebo-controlled, phase 2 trial. Lancet Diabetes Endocrinol 1, 306–316 (2013). https://doi.org:10.1016/s2213-8587(13)70065-2

45 Gottlieb, P. A. et al. Failure to preserve beta-cell function with mycophenolate mofetil and daclizumab combined therapy in patients with new-onset type 1 diabetes. Diabetes Care 33, 826–832 (2010). https://doi.org:10.2337/dc09-1349

46 Greenbaum, C. J. et al. IL-6 receptor blockade does not slow beta cell loss in new-onset type 1 diabetes. JCI Insight 6 (2021). https://doi.org:10.1172/jci.insight.150074

47 Griffin, K. J., Thompson, P. A., Gottschalk, M., Kyllo, J. H. & Rabinovitch, A. Combination therapy with sitagliptin and lansoprazole in patients with recent-onset type 1 diabetes (REPAIR-T1D): 12-month results of a multicentre, randomised, placebo-controlled, phase 2 trial. Lancet Diabetes Endocrinol 2, 710–718 (2014). https://doi.org:10.1016/s2213-8587(14)70115-9

48 Groele, L. et al. Lack of effect of Lactobacillus rhamnosus GG and Bifidobacterium lactis Bb12 on beta-cell function in children with newly diagnosed type 1 diabetes: a randomised controlled trial. BMJ Open Diabetes Res Care 9 (2021). https://doi.org:10.1136/bmjdrc-2020-001523

49 Hagopian, W. et al. Teplizumab preserves C-peptide in recent-onset type 1 diabetes: two-year results from the randomized, placebo-controlled Protégé trial. Diabetes 62, 3901–3908 (2013). https://doi.org:10.2337/db13-0236

50 Haller, M. J. et al. Low-Dose Anti-Thymocyte Globulin Preserves C-Peptide, Reduces HbA(1c), and Increases Regulatory to Conventional T-Cell Ratios in New-Onset Type 1 Diabetes: Two-Year Clinical Trial Data. Diabetes 68, 1267-1276 (2019). https://doi.org:10.2337/db19-0057

51 Haller, M. J. et al. Low-Dose Anti-Thymocyte Globulin (ATG) Preserves β-Cell Function and Improves HbA(1c) in New-Onset Type 1 Diabetes. Diabetes Care 41, 1917–1925 (2018). https://doi.org:10.2337/dc18-0494

53. Herold, K. C. et al. Teplizumab (anti-CD3 mAb) treatment preserves C-peptide responses in patients with new-onset type 1 diabetes in a randomized controlled trial: metabolic and immunologic features at baseline identify a subgroup of responders. Diabetes 62, 3766–3774 (2013). https://doi.org:10.2337/db13-0345

53 Herold, K. C. et al. Teplizumab treatment may improve C-peptide responses in participants with type 1 diabetes after the new-onset period: a randomised controlled trial. Diabetologia 56, 391–400 (2013). https://doi.org:10.1007/s00125-012-2753-4

54 Herold, K. C. et al. Increased T cell proliferative responses to islet antigens identify clinical responders to anti-CD20 monoclonal antibody (rituximab) therapy in type 1 diabetes. J Immunol 187, 1998–2005 (2011). https://doi.org:10.4049/jimmunol.1100539

55 Keymeulen, B. et al. Insulin needs after CD3-antibody therapy in new-onset type 1 diabetes. N Engl J Med 352, 2598–2608 (2005). https://doi.org:10.1056/NEJMoa043980

56 Keymeulen, B. et al. Four-year metabolic outcome of a randomised controlled CD3-antibody trial in recent-onset type 1 diabetic patients depends on their age and baseline residual beta cell mass. Diabetologia 53, 614–623 (2010). https://doi.org:10.1007/s00125-009-1644-9

57 Kumar, S. et al. A high potency multi-strain probiotic improves glycemic control in children with new-onset type 1 diabetes mellitus: A randomized, double-blind, and placebo-controlled pilot study. Pediatr Diabetes 22, 1014–1022 (2021). https://doi.org:10.1111/pedi.13244

58 Lagarde, W. H. et al. Human plasma-derived alpha(1) -proteinase inhibitor in patients with new-onset type 1 diabetes mellitus: A randomized, placebo-controlled proof-of-concept study. Pediatr Diabetes 22, 192–201 (2021). https://doi.org:10.1111/pedi.13162

59 Lebenthal, Y., et al. A Phase II, Double-Blind, Randomized, Placebo-Controlled, Multicenter Study Evaluating the Efficacy and Safety of Alpha-1 Antitrypsin (AAT) (Glassia(®)) in the Treatment of Recent-Onset Type 1 Diabetes. Int J Mol Sci 20 (2019). https://doi.org:10.3390/ijms20236032

60 Linsley, P. S. et al. Elevated T cell levels in peripheral blood predict poor clinical response following rituximab treatment in new-onset type 1 diabetes. Genes Immun 20, 293–307 (2019). https://doi.org:10.1038/s41435-018-0032-1

61 Linsley, P. S., Greenbaum, C. J., Speake, C., Long, S. A. & Dufort, M. J. B lymphocyte alterations accompany abatacept resistance in new-onset type 1 diabetes. JCI Insight 4 (2019). https://doi.org:10.1172/jci.insight.126136

62 Long, S. A. et al. Partial exhaustion of CD8 T cells and clinical response to teplizumab in new-onset type 1 diabetes. Sci Immunol 1 (2016). https://doi.org:10.1126/sciimmunol.aai7793

63 Long, S. A. et al. Remodeling T cell compartments during anti-CD3 immunotherapy of type 1 diabetes. Cell Immunol 319, 3–9 (2017). https://doi.org:10.1016/j.cellimm.2017.07.007

64 Ludvigsson, J. et al. GAD65 antigen therapy in recently diagnosed type 1 diabetes mellitus. N Engl J Med 366, 433–442 (2012). https://doi.org:10.1056/NEJMoa1107096

66. Ludvigsson, J. et al. Combined vitamin D, ibuprofen and glutamic acid decarboxylase-alum treatment in recent onset Type i diabetes: Lessons from the DIABGAD randomized pilot trial. Future Science OA 6 (2020). https://doi.org:10.2144/fsoa-2020-0078

66 Ludvigsson, J. et al. Intralymphatic Glutamic Acid Decarboxylase With Vitamin D Supplementation in Recent-Onset Type 1 Diabetes: A Double-Blind, Randomized, Placebo-Controlled Phase IIb Trial. Diabetes Care 44, 1604–1612 (2021). https://doi.org:10.2337/dc21-0318

67 Martin, S. et al. Residual beta cell function in newly diagnosed type 1 diabetes after treatment with atorvastatin: the Randomized DIATOR Trial. PLoS One 6, e17554 (2011). https://doi.org:10.1371/journal.pone.0017554

68 Moran, A. et al. Interleukin-1 antagonism in type 1 diabetes of recent onset: two multicentre, randomised, double-blind, placebo-controlled trials. Lancet 381, 1905–1915 (2013). https://doi.org:10.1016/s0140-6736(13)60023-9

69 Nafei, L. T., Kadhim, K. A., Said, A. M. & Ghani, S. H. Evaluation the effect of vitamin D3 on glycemic indices on Iraqi children with type 1 DM. International Journal of Pharmaceutical Sciences Review and Research 42, 134–143 (2017).

70 Orban, T. et al. Reduction in CD4 central memory T-cell subset in costimulation modulator abatacept-treated patients with recent-onset type 1 diabetes is associated with slower C-peptide decline. Diabetes 63, 3449–3457 (2014). https://doi.org:10.2337/db14-0047

71 Orban, T. et al. Costimulation modulation with abatacept in patients with recent-onset type 1 diabetes: follow-up 1 year after cessation of treatment. Diabetes Care 37, 1069–1075 (2014). https://doi.org:10.2337/dc13-0604

72 Orban, T. et al. Co-stimulation modulation with abatacept in patients with recent-onset type 1 diabetes: a randomised, double-blind, placebo-controlled trial. Lancet 378, 412–419 (2011). https://doi.org:10.1016/s0140-6736(11)60886-6

73 Ortqvist, E. et al. Temporary preservation of beta-cell function by diazoxide treatment in childhood type 1 diabetes. Diabetes Care 27, 2191–2197 (2004). https://doi.org:10.2337/diacare.27.9.2191

74 Pescovitz, M. D. et al. B-lymphocyte depletion with rituximab and β-cell function: two-year results. Diabetes Care 37, 453–459 (2014). https://doi.org:10.2337/dc13-0626

75 Pescovitz, M. D., et al. Rituximab, B-lymphocyte depletion, and preservation of beta-cell function. N Engl J Med 361, 2143–2152 (2009). https://doi.org:10.1056/NEJMoa0904452

76 Pitocco, D. et al. The effects of calcitriol and nicotinamide on residual pancreatic beta-cell function in patients with recent-onset Type 1 diabetes (IMDIAB XI). Diabet Med 23, 920–923 (2006). https://doi.org:10.1111/j.1464-5491.2006.01921.x

77 Pozzilli, P. et al. Randomized 52-week Phase 2 Trial of Albiglutide Versus Placebo in Adult Patients With Newly Diagnosed Type 1 Diabetes. J Clin Endocrinol Metab 105 (2020). https://doi.org:10.1210/clinem/dgaa149

78 Pozzilli, P. et al. No effect of oral insulin on residual beta-cell function in recent-onset type I diabetes (the IMDIAB VII). IMDIAB Group. Diabetologia 43, 1000–1004 (2000). https://doi.org:10.1007/s001250051482

80. Pozzilli, P. et al. Vitamin E and nicotinamide have similar effects in maintaining residual beta cell function in recent onset insulin-dependent diabetes (the, IMDIAB IV study). Eur J Endocrinol 137, 234–239 (1997). https://doi.org:10.1530/eje.0.1370234

80 Quattrin, T. et al. Golimumab and Beta-Cell Function in Youth with New-Onset Type 1 Diabetes. N Engl J Med 383, 2007–2017 (2020). https://doi.org:10.1056/NEJMoa2006136

81 Rother, K. I. et al. Effect of ingested interferon-alpha on beta-cell function in children with new-onset type 1 diabetes. Diabetes Care 32, 1250–1255 (2009). https://doi.org:10.2337/dc08-2029

82 Sherry, N. et al. Teplizumab for treatment of type 1 diabetes (Protégé study): 1- year results from a randomised, placebo-controlled trial. Lancet 378, 487–497 (2011). https://doi.org:10.1016/s0140-6736(11)60931-8

83 Strom, A. et al. Improved preservation of residual beta cell function by atorvastatin in patients with recent onset type 1 diabetes and high CRP levels (DIATOR trial). PLoS One 7, e33108 (2012). https://doi.org:10.1371/journal.pone.0033108

84 von Herrath, M. et al. Anti-interleukin-21 antibody and liraglutide for the preservation of β-cell function in adults with recent-onset type 1 diabetes: a randomised, double-blind, placebo-controlled, phase 2 trial. Lancet Diabetes Endocrinol 9, 212–224 (2021). https://doi.org:10.1016/s2213-8587(21)00019-x

85 Walter, M., Philotheou, A., Bonnici, F., Ziegler, A. G. & Jimenez, R. No effect of the altered peptide ligand NBI-6024 on beta-cell residual function and insulin needs in new-onset type 1 diabetes. Diabetes Care 32, 2036–2040 (2009). https://doi.org:10.2337/dc09-0449

86 Wherrett, D. K. et al. Antigen-based therapy with glutamic acid decarboxylase (GAD) vaccine in patients with recent-onset type 1 diabetes: a randomised double-blind trial. Lancet 378, 319–327 (2011). https://doi.org:10.1016/s0140-6736(11)60895-7

87 Diabetes Prevention Trial--Type 1 Diabetes Study, G. Effects of insulin in relatives of patients with type 1 diabetes mellitus. N Engl J Med 346, 1685-1691 (2002). https://doi.org:10.1056/NEJMoa012350

88 Herold, K. C. et al. An Anti-CD3 Antibody, Teplizumab, in Relatives at Risk for Type 1 Diabetes. N Engl J Med 381, 603-613 (2019). https://doi.org:10.1056/NEJMoa1902226

89 Elding Larsson, H., et al. Safety and efficacy of autoantigen-specific therapy with 2 doses of alum-formulated glutamate decarboxylase in children with multiple islet autoantibodies and risk for type 1 diabetes: A randomized clinical trial. Pediatr Diabetes 19, 410–419 (2018). https://doi.org:10.1111/pedi.12611

90 Palmer, J. P. et al. C-peptide is the appropriate outcome measure for type 1 diabetes clinical trials to preserve beta-cell function: report of an ADA workshop, 21-22 October 2001. Diabetes 53, 250-264 (2004). https://doi.org:10.2337/diabetes.53.1.250

91 ElSayed, N. A. et al. 2. Classification and Diagnosis of Diabetes: Standards of Care in Diabetes-2023. Diabetes Care 46, S19-S40 (2023). https://doi.org:10.2337/dc23-S002

92 Ziegler, A. G. et al. Seroconversion to multiple islet autoantibodies and risk of progression to diabetes in children. Jama 309, 2473–2479 (2013).

93 Insel, R. A. et al. Staging presymptomatic type 1 diabetes: a scientific statement of JDRF, the Endocrine Society, and the American Diabetes Association. Diabetes Care 38, 1964–1974 (2015). https://doi.org:10.2337/dc15-1419

94 Greenbaum, C. J. et al. Strength in Numbers: Opportunities for Enhancing the Development of Effective Treatments for Type 1 Diabetes-The TrialNet Experience. Diabetes 67, 1216–1225 (2018). https://doi.org:10.2337/db18-0065

95 Bundy, B. N., Krischer, J. P. & Type 1 Diabetes TrialNet Study, G. A quantitative measure of treatment response in recent-onset type 1 diabetes. Endocrinol Diabetes Metab 3, e00143 (2020). https://doi.org:10.1002/edm2.143

96 Sims, E. K. et al. Elevations in the Fasting Serum Proinsulin-to-C-Peptide Ratio Precede the Onset of Type 1 Diabetes. Diabetes Care 39, 1519–1526 (2016). https://doi.org:10.2337/dc15-2849

97 Krischer, J. P. et al. Genetic and Environmental Interactions Modify the Risk of Diabetes-Related Autoimmunity by 6 Years of Age: The TEDDY Study. Diabetes Care 40, 1194–1202 (2017). https://doi.org:10.2337/dc17-0238

98 So, M. et al. Characterising the age-dependent effects of risk factors on type 1 diabetes progression. Diabetologia 65, 684–694 (2022). https://doi.org:10.1007/s00125-021-05647-5

99 Leete, P. et al. Studies of insulin and proinsulin in pancreas and serum support the existence of aetiopathological endotypes of type 1 diabetes associated with age at diagnosis. Diabetologia 63, 1258–1267 (2020). https://doi.org:10.1007/s00125-020-05115-6

100 Carr, A. L. J. et al. Circulating C-Peptide Levels in Living Children and Young People and Pancreatic beta-Cell Loss in Pancreas Donors Across Type 1 Diabetes Disease Duration. Diabetes 71, 1591–1596 (2022). https://doi.org:10.2337/db22-0097

101 Eizirik, D. L. et al. The human pancreatic islet transcriptome: expression of candidate genes for type 1 diabetes and the impact of pro-inflammatory cytokines. PLoS Genet 8, e1002552 (2012). https://doi.org:10.1371/journal.pgen.1002552

102 Ramos-Rodriguez, M. et al. The impact of proinflammatory cytokines on the beta-cell regulatory landscape provides insights into the genetics of type 1 diabetes. Nat Genet 51, 1588–1595 (2019). https://doi.org:10.1038/s41588-019-0524-6

103 Juan-Mateu, J., Villate, O. & Eizirik, D. L. MECHANISMS IN ENDOCRINOLOGY: Alternative splicing: the new frontier in diabetes research. Eur J Endocrinol 174, R225–238 (2016). https://doi.org:10.1530/EJE-15-0916

104 Gonzalez-Duque, S. et al. Conventional and Neo-Antigenic Peptides Presented by beta Cells Are Targeted by Circulating Naive CD8+ T Cells in Type 1 Diabetic and Healthy Donors. Cell Metab (2018). https://doi.org:10.1016/j.cmet.2018.07.007

105 Kracht, M. J. et al. Autoimmunity against a defective ribosomal insulin gene product in type 1 diabetes. Nat Med 23, 501–507 (2017). https://doi.org:10.1038/nm.4289

106 Marre, M. L. et al. Modifying Enzymes Are Elicited by ER Stress, Generating Epitopes That Are Selectively Recognized by CD4(+) T Cells in Patients With Type 1 Diabetes. Diabetes 67, 1356-1368 (2018). https://doi.org:10.2337/db17-1166

107 Thompson, P. J. et al. Targeted Elimination of Senescent Beta Cells Prevents Type 1 Diabetes. Cell Metab 29, 1045–1060 e1010 (2019). https://doi.org:10.1016/j.cmet.2019.01.021

109. Sims, E. K. et al. Proinsulin Secretion Is a Persistent Feature of Type 1 Diabetes. Diabetes Care 42, 258-264 (2019). https://doi.org:10.2337/dc17-2625

109 Habib, T. et al. Dynamic Immune Phenotypes of B and T Helper Cells Mark Distinct Stages of T1D Progression. Diabetes 68, 1240–1250 (2019). https://doi.org:10.2337/db18-1081

110 Chatenoud, L., Primo, J. & Bach, J. F. CD3 antibody-induced dominant self tolerance in overtly diabetic NOD mice. J Immunol 158, 2947–2954 (1997).

111 Marzinotto, I. et al. Islet Autoantibody Standardization Program: interlaboratory comparison of insulin autoantibody assay performance in 2018 and 2020 workshops. Diabetologia 66, 897–912 (2023). https://doi.org:10.1007/s00125-023-05877-9

112 Sims, E. K. et al. Who Is Enrolling? The Path to Monitoring in Type 1 Diabetes TrialNet’s Pathway to Prevention. Diabetes Care 42, 2228-2236 (2019). https://doi.org:10.2337/dc19-0593

113 Divers, J. M.-D., E; Lawrence, JM; Isom, S; Dabelea, D; Dolan, L; Imperatore, G; Marcovina, S; Pettitt, DJ; Pihoker, C; Hamman, RF; Saydah, S; Wagenknecht, LE. Trends in Incidence of Type 1 and Type 2 Diabetes Among Youths — Selected Counties and Indian Reservations, United States, 2002–2015. . MMWR Morb Mortal Wkly Rep 69, 161–165 (2020). : http://dx.doi.org/10.15585/mmwr.mm6906a3

115. 114 Writing Committee for the Type 1 Diabetes TrialNet Oral Insulin Study, G., et al. Effect of Oral Insulin on Prevention of Diabetes in Relatives of Patients With Type 1 Diabetes: A Randomized Clinical Trial. JAMA 318, 1891–1902 (2017). https://doi.org:10.1001/jama.2017.17070

115 Hannelius, U., Beam, C. A. & Ludvigsson, J. Efficacy of GAD-alum immunotherapy associated with HLA-DR3-DQ2 in recently diagnosed type 1 diabetes. Diabetologia 63, 2177–2181 (2020). https://doi.org:10.1007/s00125-020-05227-z

116 Rigby, M. R. et al. Alefacept provides sustained clinical and immunological effects in new-onset type 1 diabetes patients. J Clin Invest 125, 3285–3296 (2015). https://doi.org:10.1172/JCI81722

117 Rigby, M. R. et al. Targeting of memory T cells with alefacept in new-onset type 1 diabetes (T1DAL study): 12 month results of a randomised, double-blind, placebo-controlled phase 2 trial. Lancet Diabetes Endocrinol 1, 284–294 (2013). https://doi.org:10.1016/S2213-8587(13)70111-6

118 Ovalle, F. et al. Verapamil and beta cell function in adults with recent-onset type 1 diabetes. Nat Med 24, 1108–1112 (2018). https://doi.org:10.1038/s41591-018-0089-4

119 Forlenza, G. P. et al. Effect of Verapamil on Pancreatic Beta Cell Function in Newly Diagnosed Pediatric Type 1 Diabetes: A Randomized Clinical Trial. JAMA (2023). https://doi.org:10.1001/jama.2023.2064

120 McVean, J. et al. Effect of Tight Glycemic Control on Pancreatic Beta Cell Function in Newly Diagnosed Pediatric Type 1 Diabetes: A Randomized Clinical Trial. JAMA (2023). https://doi.org:10.1001/jama.2023.2063

