## Supplemental for "Type 1 Diabetes Prevention: a systematic review of studies testing disease-modifying therapies and features linked to treatment response"

**Supplemental Figure 1:** PubMed/Embase Search Strategy

#1 Latent Autoimmune Diabetes in Adults [Mesh] OR "Diabetes Mellitus, Type 1 "[Mesh] OR "diabetes mellitus, type 1/prevention and control"[MeSH Terms] 81438 records

#2 ((((prevent* [Title/Abstract] OR delay [Title/Abstract] OR immunomod* [Title/Abstract] OR immune therapy [Title/Abstract] OR "disease modifying therapy" [Title/Abstract] OR Antibodies, Monoclonal, Humanized / therapeutic use* [Title/Abstract] OR C-peptide* [Title/Abstract]) OR ("autoantibodies/analysis"[MeSH Terms]))

1922455 records

#3 (randomized controlled trial[Publication Type] OR randomized controlled trial[Title/Abstract]) OR (random*[Title/Abstract] OR rct[Title/Abstract] OR randomized[Title/Abstract])
1422392 records

#4 #1 AND #2 13273 records

#5 #4 AND #3 1169 records

Filters: Human, English, 1996-present 880 records

**Supplemental Table 1. Studies in individuals with new or recent onset T1D (<1 year since diagnosis)**

| **Study ID** | **n** | **Intervention** | **Multi-center** | **Blinding** | **Primary Outcome(s)** | **Age** | **Positive?** |
| --- | --- | --- | --- | --- | --- | --- | --- |
| Ergun-Longmire 2004 ^1^ | 197 | 1 mg po insulin vs. 10 mg po insulin vs. placebo x 6-36 mo | Yes | Double | β Cell function | Both | No |
| Enander 2018 ^2^ | 54 | 48-72 hrs IV insulin vs. usual care | Yes | None | Not specified | Pediatric | No |
| Chaillous 2000 (Diabète Insuline Orale) ^3^ | 131 | 2.5 mg po human biogenetic insulin daily vs. 7.5 mg po insulin vs. placebo x 12 mo | Yes | Double | β Cell function | Both | No |
| Pozzilli 2000 ^4^ (IMDIAB VII) | 80 | 5 mg daily po insulin x 12 mo vs. placebo | Yes | Double | HbA1c; Insulin dose; β Cell function | Both | No |
| Crinò 2004 ^5^ (IMDIAB IX) | 64 | 25 mg/kg/d nicotinamide + 15 mg/kg/d vitamin E vs. 25 mg/kg/d nicotinamide x 2 yrs | Yes | Not Stated | β Cell function | Pediatric | No |
| Pitocco 2006 ^6^ (IMDIAB XI) | 70 | Calcitriol vs. Nicotinamide | Yes | None | β Cell function | Both | No |
| Pozzilli 1997 ^7^ (IMDIAB IV) | 84 | 15 mg/kg vitamin E vs. 25 mg/kg nicotinamide x 1 yr | Yes | Double | β Cell function | Both | No |
| Coutant 1998 ^8^ | 63 | 2.5mg/d linomide vs. placebo x 12 mo | No | Double | Not specified | Both | n/a |
| Allen 1999 ^9^ | 94 | 23106 colony forming units TICE BCG intradermal injection x 1 vs. placebo | Yes | Double | Incidence of remission | Pediatric | No |
| Keymeulen 2005 ^10^ | 80 | 0.22 IU/kg/day ChAglyCD3 (otelixizumab) IV x 6 days vs. placebo | Yes | Double | β Cell function | Both | Yes |
| - Keymeulen 2010 ^11^ (follow-up) | - 73 |  |  |  |  |  | Yes |
| - Demeester 2015 ^12^ (precision) | - 80 |  |  |  |  |  |  |
| Sherry 2011 ^13^ (Protégé) | 516 | 14 d full dose, 14 d low dose, or 6 d full dose IV teplizumab vs. placebo at wk 0 and wk 26 | Yes | Double | HbA1c; Insulin dose | Both | No |
| - Hagopian 2013 ^14^ (follow-up) | - 462 | - 24 mo follow-up |  |  | β Cell function |  | Yes |
| Herold 2013 ^15^ | 63 | 14 d IV teplizumab vs. placebo | Yes | Double | β Cell function | Both | Yes |
| Herold 2013 ^16^ (AbATE) | 83 | 14 d IV teplizumab, with repeat at 1 yr vs. placebo | Yes | None; Lab blinded | β Cell function | Both | Yes |
| - Long 2016 ^17^ (precision) | - 74 |  |  |  |  |  |  |
| - Long 2017 ^18^ (precision) | - 41 |  |  |  |  |  |  |
| Aronson 2014 ^19^ (DEFEND-1) | 272 | 8 d IV otelixizumab (3.1 mg) vs. placebo, outcome at 12 months | Yes | Double | β Cell function | Both | No |
| Ambery 2014 ^20^ (DEFEND-2) | 179 | 8 d course IV otelixizumab (3.1 mg) vs. placebo | Yes | Double | β Cell function | Both | No |
| Pescovitz 2009 ^21^ | 87 | IV rituximab days 1, 8, 15, and 22 vs. placebo | Yes | Double | β Cell function | Both | Yes |
| - Pescovitz 2014 ^22^ (follow-up) | - 77 |  |  |  | β Cell function at 24 months |  | - No |
| - Herold 2011 ^23^ (precision) | - 78 |  |  |  |  |  |  |
| - Linsley 2019 ^24^ (precision) | - 54 |  |  |  |  |  |  |
| Ortqvist 2004 ^25^ | 56 | 5-7.5 mg/kg/d po diazoxide vs. placebo x 24 mo | Yes | Double | β Cell function | Pediatric | Yes |
| Walter 2009 ^26^ | 188 | 0.1 mg sc inj of altered peptide ligand NBI-6024 at baseline, wk 2 and 4, and then qmo vs. placebo x 24 mo | Yes | Double | β Cell function | Both | No |
| Rother 2009 ^27^ | 128 | 5,000 units/d or 30,0000 units/d po human recombinant IFN-alpha vs. placebo x 1 yr | Yes | Double | β Cell function | Both | Yes |
| Gottlieb 2010 ^28^ | 126 | 600mg/m2/d po mycophenolate mofetil (MMF) x 2 yrs, vs. MMF+1m/kg IV Daclizumab x 2, vs. placebo | Yes | Double | β Cell function | Both | No |
| Wherrett 2011 ^29^ | 145 | 20μg sc inj GAD-alum x 3 vs. GAD-alum x 2 /alum x 1 vs. alum x 3 | Yes | Double | β Cell function | Both | No |
| Ludvigsson 2012 ^30^ | 334 | 20ug sc inj GAD-alum x 4,vs. 20ug sc GAD-alum x 2/placebo x 2 vs. placebo x 4 | Yes | Double | β Cell function | Both | No |
| Ludvigsson 2020 ^31^ (DIABGAD ) | 64 | 450 d Vit. D + 90 d Ibuprofen + 20 μg sc inj GAD-alum x 2, vs. 40 μg GAD-alum, vs. Vit D +placebo+20 μg GAD-alum, vs. placebo | Yes | Double | Not specified | Pediatric | No |
| Ludvigsson 2021 ^32^ (DIAGNODE-2) | 109 | 4mg GAD-alum intralymphatic injection monthly x 3 + oral vitamin D (2,000 IE daily for 120 days) vs. placebo | Yes | Double | β Cell function | Both | No |
| Orban 2011 ^33^ | 112 | 10 mg/kg IV abatacept x 27 over two years vs. placebo | Yes | Double | β Cell function | Both | Yes |
| - Orban 2014 ^34^ (follow-up) | - 112 | - 36 mo extended f/u (1 yr post tx cessation) |  |  |  |  |  |
| - Orban 2014 ^35^ (precision) | - 87 |  |  |  |  |  |  |
| - Cabrera 2018 ^36^ (precision) | - 74 |  |  |  |  |  |  |
| - Linsley 2019 ^37^ (precision) | - 105 |  |  |  |  |  |  |
| - Eichmann 2020 ^38^ (precision) | - 59 |  |  |  |  |  |  |
| Martin 2011 ^39^ (DIATOR) | 89 | 80 mg/d atorvastatin vs. placebo x 18 mo | Yes | Double | β Cell function | Adult | No |
| - Strom 2012 ^40^ (precision) | - 89 |  |  |  |  |  |  |
| Moran 2013 ^41^ | 69 | 2mg/kg sc injection monthly canakinumab x 12 mo vs. placebo | Yes | Double | β Cell function | Both | No |
| Moran 2013 ^41^ (AIDA) | 69 | 100 mg daily sc inj anakinra x 9 months vs. placebo | Yes | Double | β Cell function | Both | No |
| Gitelman 2013 ^42^ (START) | 58 | 6.5 mg/kg IV ATG vs. placebo | Yes | Double blinded after 3 mo | β Cell function | Both | No |
| - Gitelman 2016 ^43^ (follow-up) | - 58 | 24-mo follow up |  |  |  |  | No |
| Ataie-Jafari 2013 ^44^ | 61 | 0.25mcg- 0.5 mcg/d po Alfacalcidol vs. placebo x 6 mo | Yes | Single | β Cell function; insulin dose | Pediatric | Yes |
| Nafei 2017 ^45^ | 75 | 2000IU/day Vit. D3 vs. usual care | No | None | Not specified | Pediatric | Yes |
| Buckingham 2013 ^46^ | 68 | Hybrid closed loop using the Medtronic MiniMed system for 72-96 hrs vs. usual care | Yes | Outcomes masked | Not specified | Both | No |
| Griffin 2014 ^47^ (REPAIR-T1D) | 68 | 50-100 mg po sitagliptin + 30-60 mg po lansoprazole x12 months vs. placebo | Yes | Double | β Cell function | Both | No |
| Pozzilli 2020 ^48^ | 67 | Albiglutide sc injection 30 -50 mg weekly x 52 wks vs. placebo | Yes | Double | β Cell function | Adult | No |
| Haller 2018 ^49^ | 89 | 2.5 mg/kg IV ATG vs. ATG + 6mg sc inj pegylated GCSF q2wks x 6 vs. placebo | Yes | Double | β Cell function | Both | Yes |
| - Haller 2019 ^50^ (follow-up) | - 89 | - 24??? mo extended f/u |  |  |  |  | Yes |
| Lebenthal 2019 ^51^ | 70 | 60 mg/kg IV Alpha-1 Antitrypsin (Glassia) x 22 (52 wks), vs. 120 mg/kg x 22 vs. placebo | Yes | Double | β Cell function | Both | No |
| Quattrin 2020 ^52^ (T1GER) | 84 | Golimumab 0, 2 wk induction then q2wk maintenance inj vs. placebo x 52 wks | Yes | Double | β Cell function | Both | Yes |
| vonHerrath 2021 ^53^ | 308 | 12 mg/kg monoclonal anti-IL-21 antibody q6 weeks vs. daily liraglutide sc inj vs. anti-IL-21 + liraglutide vs. placebo x 54 wks | Yes | Double | β Cell function | Adult | Yes |
| Lagarde 2021 ^54^ | 76 | 90 or 180 mg/kg IV human-derived alpha1-proteinase inhibitor wkly x 13 wks vs. 26 wks vs. placebo | Yes | Partial blinding | β Cell function | Both | No |
| Kumar 2021 ^55^ | 96 | 3 mo high dose po multi-strain probiotic vs. placebo | No | Double | HbA1c | Pediatric | Yes |
| Groele 2021 ^56^ | 96 | 10^9 colony-forming units/day po L. rhamnosus GG and B. lactis Bb12 vs. placebo x 6 mo | n/a | Double | β Cell function | Pediatric | No |
| Gitelman 2021 ^57^ | 67 | 400 mg po daily imatinib mesylate x 26 weeks vs. placebo | Yes | Double | β Cell function | Adult | Yes |
| Greenbaum 2021 ^58^ (EXTEND) | 163 | 8 mg/kg IV Tocilizumab monthly x 7 vs. placebo | Yes | Double | β Cell function | Both | No |
| Diggins 2021 (T1DAL) ^59^ (precision) | 26/49 | 15 mg alefacept IM qwk x 12; 12 wk off, qwk x 12 vs. placebo | Yes | Double | β Cell function | Both | No |
| Christie 2002 ^60^ (Precision) | 97/188 | Cyclosporin vs. placebo x12 mo | Yes | Double | n/a | Both | n/a |

Follow-up or precision studies describing a randomized trial that is already included in the table are listed as bulleted subheadings.

Po – per oral/orally, IV – intravenous, IMDIAB –nicotinamide in recent-onset IDDM study , HbA1c - hemoglobin A1c, TICE BCG – Tice Bacillus Calmette-Guerin, ChAglyCD3 – Otelexizumab, AbATE - Autoimmunity-Blocking Antibody for Tolerance trial, DEFEND - Durable Response Therapy Evaluation for Early or New-Onset Type 1 Diabetes, Sc – subcutaneous, Inj – injection, Qmo – every month, MMF – Mycophenolate Mofetil, GAD-Alum – Glutamic acid decarboxylase – alum, , f/u – follow-up, tx – treatment, DIATOR – Diabetes Intervention with Atorvastatin, AIDA – Anti-Interleukin-1 in Diabetes Action, START - Study of Antithymocyte Globulin for Treatment of New-onset T1DM, ATG – Antithymocyte Globulin, GCSF - Granulocyte Colony-Stimulating Factor, Q2wks – every 2 weeks

T1GER - SIMPONI to Arrest β-cell Loss in Type 1 Diabetes, Q6 – every 6, n/a – not applicable, EXTEND - Tocilizumab (TCZ) in New-onset Type 1 Diabetes, T1DAL - Inducing Remission in Type 1 Diabetes With Alefacept, IM – intramuscular, Qwk – every week

**Supplemental Table 2. Metabolic outcomes from studies in individuals with new or recent onset T1D.**

| **Study ID** | **n** | **Intervention** | **C-peptide AUC Analysis** | **Results (p value vs. control) (units in nmol/L or pmol/mL unless stated)** |
| --- | --- | --- | --- | --- |
| Enander ^2^ 2018 | 54 | IV insulin vs. usual care | Mean ± SD of 2 hr MMTT AUC | Usual Care: 23.9 ± 40.6 nmol/L*min  IV insulin: 18.26 ± 16.45 (ns) |
| Sherry 2011 ^13^(Protégé)* | 516 | 14 d full dose x 2, 14 d low dose x 2, or 6 d full dose x 2 IV teplizumab vs. placebo | 1 yr median (IQR) change from baseline in 4 hr MMTT AUC | Placebo: −0·14 (−0·30, 0·02) nmol/L*min  14 d full dose: -0.06 (-0.25, 0.12), p =0.0486;  14 d low dose: −0·13 (−0·33, 0·01);  6 d full dose: -0.08 (-0.31, 0.11) |
| Hagopian 2013 ^14^(f/u)* | 516 |  | 2 yr mean change from baseline in 4-hr MMTT AUC adjusted for age group and baseline value | Placebo:-0.191;  14 d full dose: -0.136, p=0.027;  14 d low dose: -0.198, p=0.968;  6 d full dose: -0.174, p=0.312 |
| Herold 2013*^15^ | 63 | IV teplizumab vs. placebo | 12 mo mean (95% CI) 4 hr MMTT AUC | Placebo: 0.37 (0.32, 0.42);  Teplizumab: 0.45 (0.40, 0.51), p=0.03 |
| Herold 2013 ^16^(AbATE)* | 83 | IV teplizumab, with repeat course at 1 yr vs. placebo | 24 mo mean (95% CI) change in 4 hr MMTT ln(AUC + 1) adjusted for baseline value | Placebo: -0.46 (-0.57, -0.35);  Teplizumab: -0.28 (-0.36,-0.20), p=0.002 |
| Aronson 2014 (DEFEND-1) ^19^ | 272 | IV otelixizumab vs. placebo | 12 mo change in 2 hr MMTT AUC | Placebo: -0.2 ± 0.037;  Otelixizumab: 0.025 ± 0.025 p=0.58 |
| Ambery 2014 (DEFEND-2)^20^ | 179 | IV otelixizumab vs. placebo | Difference in 12 mo change in 2 hr MMTT adjusted for age, continent, and baseline value | –0.09 (95% CI –0.17 to 0; P = 0.051) |
| Pescovitz ^21^2009* | 87 | IV rituximab vs. placebo | 12 mo mean (95% CI) 2 hr MMTT loge([mean AUC]+1) adjusted for age and sex. | Placebo: 0.47 (0.39, 0.55)  Rituximab: 0.56 (0.50, 0.63), p=0.009 |
| Pescovitz 2014 ^22^(F/u) | 77 |  | 24 mo mean (95% CI) 2 hr MMTT loge([mean AUC]+1) adjusted for age and sex. | Placebo: 0.336 (0.245‚ 0.433)  Rituximab: 0.398( 0.326‚ 0.473), p=0.15 |
| Walter 2009^26^ | 188 | NBI-6024 vs. placebo | 24 mo mean ± SD 2 hr MMTT AUC | Placebo: 50 ± 45 pmol x min/ml;  NBI-6024: 57 ± 71; p=0.5 |
| Rother 2009*^27^ | 128 | Human recombinant interferon-$\alpha$(hrIFN- $\alpha$) vs. placebo | Mean ± SD % 2 hr MMTT AUC loss from 0-12 months | Placebo: 56±29 %;  5000 units hrIFN-$\alpha$: 29±54, p=0.017;  30,000 units hrIFN-$\alpha$: 48±35, p=0.599 |
| Gottlieb 2010 ^28^ | 126 | Mycophenolate mofetil (MMF), vs. MMF+ Daclizumab (DZB), vs. placebo | 2 yr geometric mean (95%CI) 2 hr MMTT AUC | Placebo: 0.27 (0.18 ‚0.37);  MMF: 0.25 (0.14 ‚ 0.37) p=0.41;  MMF +DZB: 0.28 (0.19 ‚ 0.37) p=0.47 |
| Wherrett 2011 ^29^ | 145 | GAD-alum x 3 vs. GAD-alum x2/ alum x 1 vs. alum x 3 | Ratio (95% CI) of population mean from first 2 hr AUC of 4 hr MMTT adjusted for age, sex, and baseline C-peptide | GAD-alum ×3: 0.998 (0.779, 1.22), p = 0.98;  GAD-alum ×2/alum ×1: 0.926 (0.720, 1.13), p = 0.50) |
| Ludvigsson 2012 ^30^ | 334 | GAD-alum x 4,vs. GAD-alum x 2 placebo x 2 vs. placebo | 15 mo. mean estimated treatment ratio (95%CI) of change in 2 hr MMTT C-peptide AUC. | GAD x 4: 1.18 (0.955 - 1.458) p=0.13; GADx2/Placebo x 2: 1.149 (0.929 - 1.421), p=0.2 |
| Ludvigsson 2021 (DIAGNODE-2)^32^ | 109 | Intralymphatic GAD-alum + vitamin D (vs. placebo | Mean (95%CI) treatment effect ratio from 2 hr MMTT AUC | 1.091 (0.845-1.408); p=0.5009 |
| Orban 2011*^33^ | 112 | Abatacept vs. placebo | 2 year geometric mean (95%CI) of 2 hr MMTT AUC adjusted for age, sex, and baseline value | Placebo: 0.266 (0.171, 0.368)  Abatacept: 0·375 (0.290, 0.465) (p=0.0029) |
| Orban 2014 ^34^(f/u)* | 112 |  | 36 mo population mean (95%CI) MMTT 2-h AUC, adjusted for age, sex, and baseline value | Placebo: 0.141 (0.071‚0.215);  Abatacept: 0.217 (0.168‚0.268), p=0.046 |
| Moran 2013^41^ | 69 | Canakinumab vs. placebo | 12 mo mean (95%CI) difference in 2 hr MMTT AUC vs. placebo | 0.01 (−0.11 to 0.14), p=0.86) |
| Moran 2013 ^41^(AIDA) | 69 | Anakinra vs. placebo | 9 mo mean (95%CI) difference in 2 hr MMTT AUC vs. placebo | 0.02 (−0.09 to 0.15), p=0.71 |
| Gitelman 2013 ^42^(START) | 58 | ATG vs. placebo | 12 mo mean change (95%CI) in 2 hr MMTT AUC | Placebo: −0.239 (−0.361,−0.118)  ATG: −0.195 (−0.292, −0.098), p=0.591 |
| Gitelman ^43^2016 (f/u) | 58 |  | 24 mo change in the mean (95% CI) 2 hr C-peptide AUC from 4 hr MMTT, adjusted for baseline value | Placebo: -0.32 (-0.473,0.174);  ATG: -0.27 (-0.373,0.171), p=0.38 |
| Buckingham 2013^46^ | 68 | Hybrid closed loop (HCL) vs. usual care | Geometric mean (95%CI) 2 hr MMTT AUC | Usual care: 0.52 (0.32‚0.75);  HCL: 0.43 (0.34‚0.52) p=0.49 |
| Griffin 2014 (REPAIR-T1D)^47^ | 68 | Sitagliptin + lansoprazole vs. placebo | 12 mo mean change (95%CI) in 2 hr MMTT AUC | Placebo: −253 (−383,−123) ;  Sitagliptin+Lansoprazole: −229 (−316,−142), p=0.77 |
| Pozzilli 2020^48^ | 67 | Albiglutide sc injection 30 -50 mg weekly x 52 wks vs. placebo | 52 wk Difference in least squares means (95%CI) vs placebo for change in 2 hr MMTT AUC | 0.04 (−0.13, 0.20) , p=0.6505 |
| Haller 2018*^49^ | 89 | Low-dose ATG vs. ATG + GCSF vs. placebo | 12 mo geometric-like means (95%CI) 2 hr AUC of 4 hr MMTT adjusted for for sex, age, and baseline value. | Placebo: 0.406 (0.324, 0.494)  ATG: 0.646 (0.547, 0.750), p = 0.0003  ATG/GCSF: 0.528 (0.435, 0.627), p = 0.031) |
| Haller 2019 (f/u)*^50^ | 89 |  | 24 mo geometric-like means (95%CI) 2 hr AUC of 4 hr MMTT adjusted for for sex, age, and baseline value. | Placebo: 0.253 (0.177, 0.334)  ATG: 0.5 (0.412, 0.594) p=0<0.001  ATG+GCSF: 0.36 (0.281, 0.445)p=0.032 |
| Lebenthal 2019^51^ | 70 | 60 mg/kg IV Alpha-1 Antitrypsin (A1AT) x vs. 120 mg/kg x 22 vs. placebo | 52 wk change in 2 hr MMTT AUC | Placebo: -0.34  60mg/kg A1AT:- 0.55 p=0.677  120mg/kg A1AT:- 0.29 p=0.822 |
| Quattrin 2020 (T1GER)*^52^ | 84 | Golimumab vs. placebo | Mean ± SD 52 wk 4 hr MMTT AUC | Placebo: 0.43±0.39;  Golimumab: 0.64±0.42; p<0.001 |
| vonHerrath 2021*^53^ | 308 | Anti-IL-21 vs. liraglutide vs. combination vs. placebo | 54 wk estimated mean (95%CI) treatment ratio based on change in 4 hr MMTT AUC | IL-21: 1.23 (0.97–1.57), p=0·093  Liraglutide: 1.12 (0.87–1.42), p=0.38  Combination: 1.48 (1.16–1.89), p=0.0017 |
| Groele 2021^56^ | 96 | L. rhamnosus GG and B. lactis Bb12 vs. placebo | 6 mo median (IQR) 2 hr MMTT AUC | Placebo: 3.30 (2.14; 4.56) ng/mL  Treatment: 3.38 (2.24; 4.52), p=0.993 |
| Gitelman 2021*^57^ | 67 | Imatinib mesylate x 26 weeks vs. placebo | Mean difference (90% CI) 2 hr AUC from 4 hr MMTT adjusted for sex, baseline age, and baseline value | 0.095 (–0.003 to 0.191), p=0.048 |
| Greenbaum 2021 ^58^(EXTEND) | 81 pediatric | Tocilizumab vs. placebo | Wk 52 mean (95% CI) change in 2 hr MMTT AUC | Placebo: 0.391 (0.47,0.31)  Tocilizumab: 0.33 (0.39,0.28), p=0.277 |

*Studies with significant differences between a treatment group and placebo.

IV – intravenous, MMTT – mixed meal tolerance test, AUC – Area Under the Curve, f/u – follow-up, AbATE - Autoimmunity-Blocking Antibody for Tolerance trial, DEFEND - Durable Response Therapy Evaluation for Early or New-Onset Type 1 Diabetes, HrIFN-α – Human recombinant interferon-alpha, MMF – Mycophenolate mofetil, DZB – Daclizumab, GAD-Alum – Glutamic acid decarboxylase – alum, AIDA – Anti-Interleukin-1 in Diabetes Action, START - Study of Antithymocyte Globulin for Treatment of New-onset T1DM, HCL – Hybrid close Loop, Sc – subcutaneous , ATG – Antithymocyte Globulin, GCSF - Granulocyte Colony-Stimulating Factor, A1AT – Alpha-1 Antitrypsin , T1GER - SIMPONI to Arrest β-cell Loss in Type 1 Diabetes, EXTEND - Tocilizumab (TCZ) in New-onset Type 1 Diabetes

**Supplemental Table 3. Papers with Precision Analyses**

| **Study ID** | **Pre-specified?** | **Sub-group #** | **Multiple comp. corrected?** | **Features used to define subgroups** | **Smallest sample size** | **Outcome** | **Summary** |
| --- | --- | --- | --- | --- | --- | --- | --- |
| **Prevention** |  |  |  |  |  |  |  |
| Knip 2018^61^ | Both | 5 | No | Age; Sex; Family History of T1D; Specific HLA genotype; Study site/geographic location | 2 | Time to T1D | No relationships with tx response to extensively hydrolyzed casein formula identified |
| Näntö-Salonen 2008^62^ | Not stated | 4 | No | Age; Aab #; Specific Aab; $\beta$ cell function measure | 29 | Time to T1D | No relationships with intranasal insulin tx response identified |
| Gale 2004^63^ | Pre-specified | 5 | No | Age; Sex; Aab #; Dysglycemia/AGT; $\beta$ cell function measure | 11 | Time to T1D | No relationships with nicotinamide tx response identified |
| Skyler 2002^64^ | Pre-specified | 2 | No | Dysglycemia/AGT | 67 | Time to T1D | No relationships with parenteral tx response identified |
| Skyler 2005^65^ | Not stated | 1 | n/a | Specific Aab | 130 | Time to T1D | Among participants with higher IAA titer, oral insulin tx associated with reduced risk of progression. |
| Vehik 2011^66^ |  | 1 | n/a | Specific Aab | 130 | Time to T1D | In 75% of original participants with median 9.1 yrs f/u, participants with higher IAA titer maintained tx effect until cessation of therapy, when effect dissipated. |
| Krischer 2017^67^ | Pre-specified | 3 | No | Aab #; Specific Aab; $\beta$ cell function measure | 55 | Time to T1D | IAA+ Participants with ICA+ or GADA and IA2A+ with low FPIR with significant tx response to oral insulin. No significant response in high FPIR group or if ICA+ and GADA or IA2A+. |
| Elding Larsson 2018^68^ | Pre-specified | 3 | No | Sex; Aab #; Dysglycemia/AGT | 7 | Time to T1D | No relationships with GAD sc inj tx response identified |
| Herold 2019^69^ | Pre-specified | 12 | No | Age; Sex; BMI; Specific HLA genotype; Specific Aab; $\beta$ cell function measure; glucose | 8 | Time to T1D | Significant effect of teplizumab vs placebo if: female, BMI >median, GADA+ positive or mIAA+, ICA-, ZnT8A-, or IA2A-, DR3-, DR4+, glucose> median, C-peptide AUC< median. |
| **Prevention Precision** |  |  |  |  |  |  |  |
| Butty 2008^70^ | Post-hoc | 6 | No | Specific HLA genotype, other genetic risk feature | 5 | Time to T1D | Enhanced effect of oral insulin in those with 1 but not 2 alleles for INS-23A SNP. |
| Sosenko 2020^71^ | Post-hoc | 1 | n/a | Diabetes progression risk score (DPTRS) | 37 | C-peptide measure; Time to T1D | For those with DPTRS >=6.75 oral insulin showed significant effect in DPT-1 and in combined data from TN and DPT-1 oral insulin studies. |
| **New Onset** |  |  |  |  |  |  |  |
| Greenbaum 2021^58^ | Both | 2 | No | Age; genetic risk feature | 5 | C-peptide measure | No relationships with tocilizumab tx response identified |
| Wherrett 2011^29^ | Both | 9 | No | Age; Sex; Specific HLA genotype; Specific Aab; $\beta$ cell function measure; HbA1c | not stated | C-peptide measure | No relationships with GAD sc x treattment response identified |
| Pescovitz 2009^21^ | Not stated | 7 | No | Age; Sex; Specific HLA genotype; Aab #; $\beta$ cell function measure; Insulin dose/regimen; HbA1c | 11 | C-peptide measure | No relationships with rituximab tx response identified |
| Martin 2011^39^ | Not stated | 7 | No | Age; BMI; Aab #; $\beta$ cell function measure; Study site/geographic location | n/a | C-peptide measure | No relationships with atorvastatin tx response identified |
| Lebenthal 2019^51^ | Both | 2 | No | Age; $\beta$ cell function measure | 1 (6% of 20 in placebo group for responder analysis) | C-peptide measure; HbA1c; insulin dose | No relationships with Alpha-1 Antitrypsin tx response identified |
| Griffin 2014^47^ | Pre-specified | 5 | No | Age; Sex; $\beta$ cell function measure; Insulin dose or regimen; HbA1c | n/a | C-peptide measure | No relationships with Sitagliptin + lansoprazole tx response identified |
| Gottlieb 2010^28^ | Not stated | 6 | No | Age; Sex; Aab #;; $\beta$ cell function measure; Insulin dose/regimen; HbA1c | 10 | C-peptide measure | No relationships with Mycophenolate mofetil (MMF) +/- Daclizumab (DZB) tx response identified |
| Chaillous 2000^3^ | Not stated | 2 | No | Age; Measure of $\beta$ cell function | n/a | C-peptide measure | No relationships with oral human biogenetic insulin tx response identified |
| Buckingham 2013^46^ | Not stated | 5 | No | Age; Sex; $\beta$ cell function measure; HbA1c; DKA at dx | 4 | C-peptide measure | No relationships with hybrid closed loop tx response identified |
| Aronson 2014^19^ | Pre-specified | 10 | No | Age; Sex; BMI; Aab #; Specific Aab; $\beta$ cell function measure; Study site/geographic location; Insulin dose or regimen; HbA1c | n/a | C-peptide measure | No relationships with Otelixizumab tx response identified |
| Rother 2009^27^ | Post-hoc | 1 | n/a | Age | 41 | C-peptide measure | No relationships with oral interferon alpha tx response identified |
| Pozzilli 2000^4^ | Not stated | 1 | n/a | Age |  | C-peptide measure; time to Aab+ | No relationships with oral insulin tx response identified |
| Sherry 2011^13^ | Pre-specified | 3 | No | Age; Study site/geographic location; Duration of dx | 31 | HbA1c | For 14-day full dose teplizumab, % participants with A1c <7% and lower insulin doses higher in 8-11 year olds, US participants, and participants randomized within 6 wks of dx |
| Hagopian 2013^72^ | Pre-specified | 6 | No | Age; $\beta$ cell function measure; Insulin dose/regimen; HbA1c; Study site/geographic location; Duration of dx | 31 | C-peptide measure | For 14-day full dose teplizumab, 2-yr adjusted mean change in C-peptide AUC showed tx effect in participants in US, randomized in 6 wks from dx, or with baseline A1c<7.5%, insulin dose<0.4 u/kg/day, C-peptide >0.65 or >0.2, or if in the 8-17 yr old age category |
| Orban 2011^33^ | Pre-specified | 7 | No | Age; Sex; Specific HLA genotype; $\beta$ cell function measure; Insulin dose/regimen; HbA1c; race | n/a | C-peptide measure | DR3+ participants with better ratio of abatacept tx effect while nonwhite participants with worse ratio of tx effect. |
| Orban 2014^34^ | Pre-specified | 8 | Yes | Age; Sex; Race; $\beta$ cell function measure; Insulin dose/regimen; HbA1c; Specific HLA genotype; | 3 | C-peptide measure | Significant impact of white race and DR3+ status to improve 3-yr C-peptide AUC ratio of tx effect for abatacept vs. placebo, although race effect may be spurious due to small sample size (n=3 in placebo group). |
| Moran 2013^41^* | Pre-specified | 9 | No | Age; Sex; BMI; Specific HLA genotype; $\beta$ cell function measure; Insulin dose or regimen; ethnicity, T1D duration | 11 | C-peptide measure | Participants with lower tertile of baseline C peptide in the canakinumab-treated group had significantly lower C-peptide concentrations at 1 year |
| Moran 2013^41^ | Pre-specified | 9 | No | Age; Sex; BMI; Specific HLA genotype; $\beta$ cell function measure; Insulin dose/regimen; ethnicity, diabetes duration | 11 | C-peptide measure | No relationships with anakinra tx response identified |
| Ludvigsson 2012^30^ | Pre-specified | 13 | No | Age; Sex; BMI; Specific HLA genotype; Specific Aab #;; $\beta$ cell function measure; Study site/geographic location; Insulin dose or regimen; HbA1c; country, days since dx, pubertal stage | n/a | C-peptide measure | Participants who were male (all regimens), had baseline daily insulin dose of 0.398-0.605 IU/kg (all regimens), from non-Nordic European countries (4-dose regimen), or had baseline Tanner pubertal stage of 2 or 3 (4 dose regimen) had higher and significant estimated tx ratios |
| Herold 2013 ^15^ | Post-hoc | 2 | No | Age; HbA1c | 7 | C-peptide measure | Improved teplizumab tx response in participants who were younger and baseline A1c< 6.5%. |
| Herold 2013 ^16^ | Post-hoc | 20 | No | Age; Sex; BMI; Specific Aab #; $\beta$ cell function measure; Immune cell phenotype; Insulin dose/regimen; HbA1c | 18 | C-peptide measure | Clinical responders to teplizumab with lower baseline A1c and insulin use; Baseline CCR4+ naive CCR6+ naive CCR4+ memory CD4+ T cells, CCR4+ naive or IFN-g+ CD8+ T cells higher in nonresponders; Baseline effector memory and CD38+ terminally differentiated CD8+ T cells lower in nonresponders. |
| Gitelman 2021^57^ | Post-hoc |  | No | Age; $\beta$ cell function measure | n/a | C-peptide measure | Lower baseline Cpeptide AUC associated with better response to imatinib mesylate |
| Pozzilli ^7^1997 | Post-hoc | 1 | n/a | Age | 15 | C-peptide measure; HbA1c; insulin dose | The insulin dose required to reach the same metabolic control (based on HbA1c) in participants <15 yrs was higher in Vit E-treated vs. nicotinamide-treated participants. |
| Ludvigsson 2021 ^32^ | Pre-specified | 1 | n/a | Specific HLA genotype | 19 | C-peptide measure | HLA DR3-DQ2+ participants showed greater tx effect of GAD-alum intralymphatic inj + oral vitamin D. |
| Keymeulen 2005^10^ | Not stated | 1 | n/a | $\beta$ cell function measure | 16 | C-peptide measure | Increase in insulin dose over follow up did not occur among participants treated with ChAglyCD3 with higher baseline glucose-clamp C-peptide release ($\geq$P50) |
| Gitelman 2013^42^ | Post-hoc | 1 | n/a | Age | 8 | C-peptide measure | No relationships with high dose ATG tx response identified. |
| Gitelman 2016^43^ | Post-hoc | 1 | n/a | Age | 20 | C-peptide measure; HbA1c; insulin dose | Older age group (22-35 years) had more “responders: based on C-peptide AUC and showed significant impact of high dose ATG on C-peptide vs. placebo. |
| Ergun-Longmire 2004^1^ | Not stated | 1 | n/a | Age | n/a | C-peptide measure | Significant benefit of 1 mg and 10mg of oral insulin among subjects >=20 yrs. In patients diagnosed before 20 yrs, 1 mg dose was ineffective, and 10 mg dose accelerated C-peptide loss. |
| Crinò 2004^5^ | Not stated | 1 | n/a | Age | 23 | C-peptide measure; HbA1c; insulin dose | For <9 yrs: at 6 months the nicotinamide (NA) +vitamin E group showed significantly higher C-peptide. For >9 yrs: NA alone showed higher C-peptide at 6 months and 9 months. |
| Coutant 1998^8^ | Post-hoc | 1 | n/a | $\beta$ cell function measure | 40 | C-peptide measure; HbA1c; insulin dose | Linomide tx associated with higher C-peptide in group with >0.1 pmol/L baseline C-peptide. |
| Ataie-Jafari 2013^44^ | Not stated | 1 | n/a | Sex | 7 | C-peptide measure; daily insulin dose | Males treated with alfacalcidol had improved fasting C-peptide and lower insulin doses by end of study vs. no improvement in females. |
| Allen 1999^9^ | Post-hoc | 1 | n/a subgroup | Age | 34 | C-peptide measure | Fasting and stimulated C-peptide lower at all time points in <10 yr group; rate of stimulated C-peptide loss more rapid in <10 yr gr |
| vonHerrath 2021^53^ | Pre-specified | 1 | n/a | $\beta$ cell function measure | 27 | C-peptide measure | Participants with baseline C-peptide > 0.6 nmol/L showed no effect of combination tx with anti-IL-21 and liraglutide. |
| Walter 2009 ^26^ | Not stated | 3 | No | Age; Sex; ethnicity | n/a | C-peptide measure | No relationships with NBI-6024 tx response identified. |
| Keymeulen 2010 ^11^ | Post-hoc | 2 | Yes | Age; $\beta$ cell function measure | 9 | C-peptide measure; HbA1c; insulin dose | ChAglyCD3-treated subgroup with initial C-peptide release $\geq$50th percentile needed lower insulin doses than the corresponding placebo subgroup, but no difference in <50th percentile group; In younger subgroup tx decreased insulin doses and metabolic control vs. placebo at months 24, 36 and 48; In the older subgroup, effect on mean insulin doses only significant and only at 24 months. |
| **New-onset Precision** |  |  |  |  |  |  |  |
| Christie 2002 ^60^ | Not stated | 5 | No | Specific Aab #; | 23 for ia2a but numbers not listed for other aabs | C-peptide measure; insulin independence; time in remission | 1) Insulin doses lower, stimulated C-peptide was higher, remission rates increased and rate of recurrence lower in cyclosporin treated IA-2A negative group  2) Cyclosporin tx decreased insulin dose and had a positive effect on stimulated C-peptide in GAD+IA-2A+, GAD-IA-2A-, and GAD+IA-2A+ groups, but had minimal effect on the GAD- IA-2A+ group. |
| Linsley 2019 ^37^ | Not stated | tested many gene modules | No | Immune cell phenotype; Other: Gene expression modules reflecting immunotypes based on whole blood RNA sequencing | n/a | C-peptide measure | A transient increase in activated B cells, reprogrammed costimulatory ligand gene expression, and reduced inhibition of anti-insulin antibodies immunotype was associated with resistance to abatacept tx; responders to drug were more likely to be older than median age. |
| Strom 2012 ^40^ | Not stated | 8 | No | Age; Sex; BMI; $\beta$ cell function measure; Immune cell phenotype; Insulin dose or regimen; Total cholesterol and CRP | n/a | C-peptide measure | Lower BMI and higher fasting baseline C-peptide associated with higher median C-peptide in placebo group but not atorvastatin group; Higher CRP in atorvastatin group but not placebo group associated with higher median C-peptide. |
| Herold 2011^23^ | Not stated | 12 | No | Immune cell phenotype | 19 | C-peptide measure | CD3+ and CD4+ cell counts were significantly higher in responders to rituximab. Nonresponders showed no change in proliferative responses to diabetes associated, islet-specific, and neuronal autoantigens over 12 mo. |
| Demeester 2015^12^ | Not stated | 7 | No | Age; Specific Aab #; $\beta$ cell function measure | 9 | C-peptide measure; | Better otelixizumab response associated with higher titers of mIAA: In the placebo group, patients with higher IAA x C-peptide levels showed rapid loss of functional $\beta$ cell mass not observed in otelixizumab group. |
| Cabrera 2018^36^ | Not stated | many transcripts analyzed | Yes | Immune cell phenotype | 13 | C-peptide measure | Higher baseline inflammatory index in placebo associated with worse C-peptide trajectory but this relationship not present in abatacept tx group, suggesting that higher baseline innate inflammation was associated with better tx response. |
| Long 2016 ^17^ | Not stated | Many immune modules | No | Immune cell phenotype | 22 | C-peptide measure | A CD8+ T cell population accumulated in teplizumab responders that phenotypically resembled exhausted T cells: expressed high levels of the transcription factor EOMES, and multiple inhibitory receptors, including TIGIT and KLRG1. |
| Long 2017^18^ | Not stated | 4 | Yes | Teplizumab anti-drug antibody (ADA) positivity | 5 | C-peptide measure; modulation of immune cell phenotype | Only 1/7 teplizumab ADA+ individuals was a clinical responder at 13 mo; ADA+ subjects failed to show CD3 modulation on both CD4+ and CD8+ T cells at the time of the 2nd course. However, after 2nd course, magnitude of CD3 modulation was similar between ADA+ and ADA – groups. |
| Diggins 2021^59^ | Not stated | 7 | Yes | Immune cell phenotype changes in association with treatment response | 6 | C-peptide measure | Greater C-peptide preservation by alefacept linked to RNAseq module of CD8+ Tcell activation- and exhaustion-associated genes. Flow cytometry data showed 2 hypoproliferative CD8+ memory cell phenotypes associated with tx response, expressing exhaustion-associated markers TIGIT and KLRG1. |
| Linsley 2019 ^24^ | Not stated | 5 | Yes | Immune cell phenotype | n/a | C-peptide measure | Whole blood RNA-seq analysis with flow cytometry f/u testing showed that a transient increase in multiple T cell populations was associated with decreased pharmacodynamic activity of rituximab, increased proliferative response to islet antigens, and rapid C-peptide loss. |
| Orban 2014 ^35^ | Pre-specified | 12 | Yes. | Immune cell phenotype | n/a | C-peptide measure; Changes in immune cell subset frequencies | Placebo-treated participants with an increase in central memor CD4 T cells showed subsequent C-peptide decline, but this effect was abrogated by abatacept tx. Abatacept tx resulted in slower C-peptide loss in association with central memory CD4 T cell contraction and naïve CD4 T cell expansion. |
| Eichmann 2020^38^ | Not stated | many subsets | No. | Immune cell phenotype | n/a | C-peptide measure | No relationship with abatacept tx response identified |

*Moran 2013 paper includes analyses from 2 trials

Abbreviations: tx- treatment; dx- diagnosis; BMI body mass index; Aab- islet autoantibody; ZnT8- zinc transporter 8 autoantibody; GADA- glutamic acid decarboxylase autantibody; IAA- insulin autoantibody; ICA- islet cell autoantibody; IA2A- islet antigen 2 autoantibody

T1D – type 1 diabetes, HLA - Human Leukocyte Antigens, Tx – treatment, Aab – autoantibody, AGT – Abnormal glucose tolerance, n/a – not applicable, IAA – Insulin autoantibody, f/u – follow-up, ICA – Islet cell autoantibody, GADA – glutamic acid decarboxylase antibody, IA2A – insulinoma-associated protein 2 autoantibody, FPIR – first phase insulin response, Sc – subcutaneous, Inj – injection, BMI – body mass index, ZnT8A - zinc transporter-8 antibody, AUC – area under the curve, SNP – single nucleotide polymorphism, DPTRS – Diabetes Prevention Trial-Type 1 Risk Score, DPT-1 – Diabetes Prevention Trial-Type 1, TN – TrialNet, HbA1c – hemoglobin A1c, GAD Glutamic Acid Decarboxylase, MMF – Mycophenolate Mofetil, DZB – Daclizumab, DKA – Diabetes Ketoacidosis, Dx – diagnosis, ChAglyCD3 – Otelexizumab, ATG - Antithymocyte Globulin, NA – Nicotinamide, Gr – group, CRP – C-reactive protein, ADA – anti-drug antibody

**Supplemental** **References**

1	Ergun-Longmire, B. *et al.* Oral insulin therapy to prevent progression of immune-mediated (type 1) diabetes. *Ann N Y Acad Sci* **1029**, 260-277 (2004). <https://doi.org:10.1196/annals.1309.057>

2	Enander, R. *et al.* Beta cell function after intensive subcutaneous insulin therapy or intravenous insulin infusion at onset of type 1 diabetes in children without ketoacidosis. *Pediatric Diabetes* **19**, 1079-1085 (2018). <https://doi.org:10.1111/pedi.12657>

3	Chaillous, L. *et al.* Oral insulin administration and residual beta-cell function in recent-onset type 1 diabetes: a multicentre randomised controlled trial. Diabète Insuline Orale group. *Lancet* **356**, 545-549 (2000). <https://doi.org:10.1016/s0140-6736(00)02579-4>

4	Pozzilli, P. *et al.* No effect of oral insulin on residual beta-cell function in recent-onset type I diabetes (the IMDIAB VII). IMDIAB Group. *Diabetologia* **43**, 1000-1004 (2000). <https://doi.org:10.1007/s001250051482>

5	Crinò, A. *et al.* A randomized trial of nicotinamide and vitamin E in children with recent onset type 1 diabetes (IMDIAB IX). *Eur J Endocrinol* **150**, 719-724 (2004). <https://doi.org:10.1530/eje.0.1500719>

6	Pitocco, D. *et al.* The effects of calcitriol and nicotinamide on residual pancreatic beta-cell function in patients with recent-onset Type 1 diabetes (IMDIAB XI). *Diabet Med* **23**, 920-923 (2006). <https://doi.org:10.1111/j.1464-5491.2006.01921.x>

7	Pozzilli, P. *et al.* Vitamin E and nicotinamide have similar effects in maintaining residual beta cell function in recent onset insulin-dependent diabetes (the IMDIAB IV study). *Eur J Endocrinol* **137**, 234-239 (1997). <https://doi.org:10.1530/eje.0.1370234>

8	Coutant, R. *et al.* Low dose linomide in Type I juvenile diabetes of recent onset: a randomised placebo-controlled double blind trial. *Diabetologia* **41**, 1040-1046 (1998). <https://doi.org:10.1007/s001250051028>

9	Allen, H. F. *et al.* Effect of Bacillus Calmette-Guerin vaccination on new-onset type 1 diabetes. A randomized clinical study. *Diabetes Care* **22**, 1703-1707 (1999). <https://doi.org:10.2337/diacare.22.10.1703>

10	Keymeulen, B. *et al.* Insulin needs after CD3-antibody therapy in new-onset type 1 diabetes. *N Engl J Med* **352**, 2598-2608 (2005). <https://doi.org:10.1056/NEJMoa043980>

11	Keymeulen, B. *et al.* Four-year metabolic outcome of a randomised controlled CD3-antibody trial in recent-onset type 1 diabetic patients depends on their age and baseline residual beta cell mass. *Diabetologia* **53**, 614-623 (2010). <https://doi.org:10.1007/s00125-009-1644-9>

12	Demeester, S. *et al.* Preexisting insulin autoantibodies predict efficacy of otelixizumab in preserving residual β-cell function in recent-onset type 1 diabetes. *Diabetes Care* **38**, 644-651 (2015). <https://doi.org:10.2337/dc14-1575>

13	Sherry, N. *et al.* Teplizumab for treatment of type 1 diabetes (Protégé study): 1-year results from a randomised, placebo-controlled trial. *Lancet* **378**, 487-497 (2011). <https://doi.org:10.1016/s0140-6736(11)60931-8>

14	Hagopian, W. *et al.* Teplizumab preserves C-peptide in recent-onset type 1 diabetes: two-year results from the randomized, placebo-controlled Protégé trial. *Diabetes* **62**, 3901-3908 (2013). <https://doi.org:10.2337/db13-0236>

15	Herold, K. C. *et al.* Teplizumab treatment may improve C-peptide responses in participants with type 1 diabetes after the new-onset period: a randomised controlled trial. *Diabetologia* **56**, 391-400 (2013). <https://doi.org:10.1007/s00125-012-2753-4>

16	Herold, K. C. *et al.* Teplizumab (anti-CD3 mAb) treatment preserves C-peptide responses in patients with new-onset type 1 diabetes in a randomized controlled trial: metabolic and immunologic features at baseline identify a subgroup of responders. *Diabetes* **62**, 3766-3774 (2013). <https://doi.org:10.2337/db13-0345>

17	Long, S. A. *et al.* Partial exhaustion of CD8 T cells and clinical response to teplizumab in new-onset type 1 diabetes. *Sci Immunol* **1** (2016). <https://doi.org:10.1126/sciimmunol.aai7793>

18	Long, S. A. *et al.* Remodeling T cell compartments during anti-CD3 immunotherapy of type 1 diabetes. *Cell Immunol* **319**, 3-9 (2017). <https://doi.org:10.1016/j.cellimm.2017.07.007>

19	Aronson, R. *et al.* Low-dose otelixizumab anti-CD3 monoclonal antibody DEFEND-1 study: results of the randomized phase III study in recent-onset human type 1 diabetes. *Diabetes Care* **37**, 2746-2754 (2014). <https://doi.org:10.2337/dc13-0327>

20	Ambery, P. *et al.* Efficacy and safety of low-dose otelixizumab anti-CD3 monoclonal antibody in preserving C-peptide secretion in adolescent type 1 diabetes: DEFEND-2, a randomized, placebo-controlled, double-blind, multi-centre study. *Diabet Med* **31**, 399-402 (2014). <https://doi.org:10.1111/dme.12361>

21	Pescovitz, M. D. *et al.* Rituximab, B-lymphocyte depletion, and preservation of beta-cell function. *N Engl J Med* **361**, 2143-2152 (2009). <https://doi.org:10.1056/NEJMoa0904452>

22	Pescovitz, M. D. *et al.* B-lymphocyte depletion with rituximab and β-cell function: two-year results. *Diabetes Care* **37**, 453-459 (2014). <https://doi.org:10.2337/dc13-0626>

23	Herold, K. C. *et al.* Increased T cell proliferative responses to islet antigens identify clinical responders to anti-CD20 monoclonal antibody (rituximab) therapy in type 1 diabetes. *J Immunol* **187**, 1998-2005 (2011). <https://doi.org:10.4049/jimmunol.1100539>

24	Linsley, P. S. *et al.* Elevated T cell levels in peripheral blood predict poor clinical response following rituximab treatment in new-onset type 1 diabetes. *Genes Immun* **20**, 293-307 (2019). <https://doi.org:10.1038/s41435-018-0032-1>

25	Ortqvist, E. *et al.* Temporary preservation of beta-cell function by diazoxide treatment in childhood type 1 diabetes. *Diabetes Care* **27**, 2191-2197 (2004). <https://doi.org:10.2337/diacare.27.9.2191>

26	Walter, M., Philotheou, A., Bonnici, F., Ziegler, A. G. & Jimenez, R. No effect of the altered peptide ligand NBI-6024 on beta-cell residual function and insulin needs in new-onset type 1 diabetes. *Diabetes Care* **32**, 2036-2040 (2009). <https://doi.org:10.2337/dc09-0449>

27	Rother, K. I. *et al.* Effect of ingested interferon-alpha on beta-cell function in children with new-onset type 1 diabetes. *Diabetes Care* **32**, 1250-1255 (2009). <https://doi.org:10.2337/dc08-2029>

28	Gottlieb, P. A. *et al.* Failure to preserve beta-cell function with mycophenolate mofetil and daclizumab combined therapy in patients with new- onset type 1 diabetes. *Diabetes Care* **33**, 826-832 (2010). <https://doi.org:10.2337/dc09-1349>

29	Wherrett, D. K. *et al.* Antigen-based therapy with glutamic acid decarboxylase (GAD) vaccine in patients with recent-onset type 1 diabetes: a randomised double-blind trial. *Lancet* **378**, 319-327 (2011). <https://doi.org:10.1016/s0140-6736(11)60895-7>

30	Ludvigsson, J. *et al.* GAD65 antigen therapy in recently diagnosed type 1 diabetes mellitus. *N Engl J Med* **366**, 433-442 (2012). <https://doi.org:10.1056/NEJMoa1107096>

31	Ludvigsson, J. *et al.* Combined vitamin D, ibuprofen and glutamic acid decarboxylase-alum treatment in recent onset Type i diabetes: Lessons from the DIABGAD randomized pilot trial. *Future Science OA* **6** (2020). <https://doi.org:10.2144/fsoa-2020-0078>

32	Ludvigsson, J. *et al.* Intralymphatic Glutamic Acid Decarboxylase With Vitamin D Supplementation in Recent-Onset Type 1 Diabetes: A Double-Blind, Randomized, Placebo-Controlled Phase IIb Trial. *Diabetes Care* **44**, 1604-1612 (2021). <https://doi.org:10.2337/dc21-0318>

33	Orban, T. *et al.* Co-stimulation modulation with abatacept in patients with recent-onset type 1 diabetes: a randomised, double-blind, placebo-controlled trial. *Lancet* **378**, 412-419 (2011). <https://doi.org:10.1016/s0140-6736(11)60886-6>

34	Orban, T. *et al.* Costimulation modulation with abatacept in patients with recent-onset type 1 diabetes: follow-up 1 year after cessation of treatment. *Diabetes Care* **37**, 1069-1075 (2014). <https://doi.org:10.2337/dc13-0604>

35	Orban, T. *et al.* Reduction in CD4 central memory T-cell subset in costimulation modulator abatacept-treated patients with recent-onset type 1 diabetes is associated with slower C-peptide decline. *Diabetes* **63**, 3449-3457 (2014). <https://doi.org:10.2337/db14-0047>

36	Cabrera, S. M. *et al.* Innate immune activity as a predictor of persistent insulin secretion and association with responsiveness to CTLA4-Ig treatment in recent-onset type 1 diabetes. *Diabetologia* **61**, 2356-2370 (2018). <https://doi.org:10.1007/s00125-018-4708-x>

37	Linsley, P. S., Greenbaum, C. J., Speake, C., Long, S. A. & Dufort, M. J. B lymphocyte alterations accompany abatacept resistance in new-onset type 1 diabetes. *JCI Insight* **4** (2019). <https://doi.org:10.1172/jci.insight.126136>

38	Eichmann, M. *et al.* Costimulation Blockade Disrupts CD4(+) T Cell Memory Pathways and Uncouples Their Link to Decline in beta-Cell Function in Type 1 Diabetes. *J Immunol* **204**, 3129-3138 (2020). <https://doi.org:10.4049/jimmunol.1901439>

39	Martin, S. *et al.* Residual beta cell function in newly diagnosed type 1 diabetes after treatment with atorvastatin: the Randomized DIATOR Trial. *PLoS One* **6**, e17554 (2011). <https://doi.org:10.1371/journal.pone.0017554>

40	Strom, A. *et al.* Improved preservation of residual beta cell function by atorvastatin in patients with recent onset type 1 diabetes and high CRP levels (DIATOR trial). *PLoS One* **7**, e33108 (2012). <https://doi.org:10.1371/journal.pone.0033108>

41	Moran, A. *et al.* Interleukin-1 antagonism in type 1 diabetes of recent onset: two multicentre, randomised, double-blind, placebo-controlled trials. *Lancet* **381**, 1905-1915 (2013). <https://doi.org:10.1016/s0140-6736(13)60023-9>

42	Gitelman, S. E. *et al.* Antithymocyte globulin treatment for patients with recent-onset type 1 diabetes: 12-month results of a randomised, placebo-controlled, phase 2 trial. *Lancet Diabetes Endocrinol* **1**, 306-316 (2013). <https://doi.org:10.1016/s2213-8587(13)70065-2>

43	Gitelman, S. E. *et al.* Antithymocyte globulin therapy for patients with recent-onset type 1 diabetes: 2 year results of a randomised trial. *Diabetologia* **59**, 1153-1161 (2016). <https://doi.org:10.1007/s00125-016-3917-4>

44	Ataie-Jafari, A. *et al.* A randomized placebo-controlled trial of alphacalcidol on the preservation of beta cell function in children with recent onset type 1 diabetes. *Clin Nutr* **32**, 911-917 (2013). <https://doi.org:10.1016/j.clnu.2013.01.012>

45	Nafei, L. T., Kadhim, K. A., Said, A. M. & Ghani, S. H. Evaluation the effect of vitamin D3 on glycemic indices on Iraqi children with type 1 DM. *International Journal of Pharmaceutical Sciences Review and Research* **42**, 134-143 (2017).

46	Buckingham, B. *et al.* Effectiveness of early intensive therapy on β-cell preservation in type 1 diabetes. *Diabetes Care* **36**, 4030-4035 (2013). <https://doi.org:10.2337/dc13-1074>

47	Griffin, K. J., Thompson, P. A., Gottschalk, M., Kyllo, J. H. & Rabinovitch, A. Combination therapy with sitagliptin and lansoprazole in patients with recent-onset type 1 diabetes (REPAIR-T1D): 12-month results of a multicentre, randomised, placebo-controlled, phase 2 trial. *Lancet Diabetes Endocrinol* **2**, 710-718 (2014). <https://doi.org:10.1016/s2213-8587(14)70115-9>

48	Pozzilli, P. *et al.* Randomized 52-week Phase 2 Trial of Albiglutide Versus Placebo in Adult Patients With Newly Diagnosed Type 1 Diabetes. *J Clin Endocrinol Metab* **105** (2020). <https://doi.org:10.1210/clinem/dgaa149>

49	Haller, M. J. *et al.* Low-Dose Anti-Thymocyte Globulin (ATG) Preserves β-Cell Function and Improves HbA(1c) in New-Onset Type 1 Diabetes. *Diabetes Care* **41**, 1917-1925 (2018). <https://doi.org:10.2337/dc18-0494>

50	Haller, M. J. *et al.* Low-Dose Anti-Thymocyte Globulin Preserves C-Peptide, Reduces HbA(1c), and Increases Regulatory to Conventional T-Cell Ratios in New-Onset Type 1 Diabetes: Two-Year Clinical Trial Data. *Diabetes* **68**, 1267-1276 (2019). <https://doi.org:10.2337/db19-0057>

51	Lebenthal, Y. *et al.* A Phase II, Double-Blind, Randomized, Placebo-Controlled, Multicenter Study Evaluating the Efficacy and Safety of Alpha-1 Antitrypsin (AAT) (Glassia(®)) in the Treatment of Recent-Onset Type 1 Diabetes. *Int J Mol Sci* **20** (2019). <https://doi.org:10.3390/ijms20236032>

52	Quattrin, T. *et al.* Golimumab and Beta-Cell Function in Youth with New-Onset Type 1 Diabetes. *N Engl J Med* **383**, 2007-2017 (2020). <https://doi.org:10.1056/NEJMoa2006136>

53	von Herrath, M. *et al.* Anti-interleukin-21 antibody and liraglutide for the preservation of β-cell function in adults with recent-onset type 1 diabetes: a randomised, double-blind, placebo-controlled, phase 2 trial. *Lancet Diabetes Endocrinol* **9**, 212-224 (2021). <https://doi.org:10.1016/s2213-8587(21)00019-x>

54	Lagarde, W. H. *et al.* Human plasma-derived alpha(1) -proteinase inhibitor in patients with new-onset type 1 diabetes mellitus: A randomized, placebo-controlled proof-of-concept study. *Pediatr Diabetes* **22**, 192-201 (2021). <https://doi.org:10.1111/pedi.13162>

55	Kumar, S. *et al.* A high potency multi-strain probiotic improves glycemic control in children with new-onset type 1 diabetes mellitus: A randomized, double-blind, and placebo-controlled pilot study. *Pediatr Diabetes* **22**, 1014-1022 (2021). <https://doi.org:10.1111/pedi.13244>

56	Groele, L. *et al.* Lack of effect of Lactobacillus rhamnosus GG and Bifidobacterium lactis Bb12 on beta-cell function in children with newly diagnosed type 1 diabetes: a randomised controlled trial. *BMJ Open Diabetes Res Care* **9** (2021). <https://doi.org:10.1136/bmjdrc-2020-001523>

57	Gitelman, S. E. *et al.* Imatinib therapy for patients with recent-onset type 1 diabetes: a multicentre, randomised, double-blind, placebo-controlled, phase 2 trial. *Lancet Diabetes Endocrinol* **9**, 502-514 (2021). <https://doi.org:10.1016/s2213-8587(21)00139-x>

58	Greenbaum, C. J. *et al.* IL-6 receptor blockade does not slow beta cell loss in new-onset type 1 diabetes. *JCI Insight* **6** (2021). <https://doi.org:10.1172/jci.insight.150074>

59	Diggins, K. E. *et al.* Exhausted-like CD8+ T cell phenotypes linked to C-peptide preservation in alefacept-treated T1D subjects. *JCI Insight* **6** (2021). <https://doi.org:10.1172/jci.insight.142680>

60	Christie, M. R., Mølvig, J., Hawkes, C. J., Carstensen, B. & Mandrup-Poulsen, T. IA-2 antibody-negative status predicts remission and recovery of C-peptide levels in type 1 diabetic patients treated with cyclosporin. *Diabetes Care* **25**, 1192-1197 (2002). <https://doi.org:10.2337/diacare.25.7.1192>

61	Knip, M. *et al.* Effect of Hydrolyzed Infant Formula vs Conventional Formula on Risk of Type 1 Diabetes. *JAMA* **319**, 38 (2018). <https://doi.org:10.1001/jama.2017.19826>

62	Näntö-Salonen, K. *et al.* Nasal insulin to prevent type 1 diabetes in children with HLA genotypes and autoantibodies conferring increased risk of disease: a double-blind, randomised controlled trial. *Lancet* **372**, 1746-1755 (2008). <https://doi.org:10.1016/s0140-6736(08)61309-4>

63	Gale, E. A., Bingley, P. J., Emmett, C. L. & Collier, T. European Nicotinamide Diabetes Intervention Trial (ENDIT): a randomised controlled trial of intervention before the onset of type 1 diabetes. *Lancet* **363**, 925-931 (2004). <https://doi.org:10.1016/s0140-6736(04)15786-3>

64	Effects of insulin in relatives of patients with type 1 diabetes mellitus. *N Engl J Med* **346**, 1685-1691 (2002). <https://doi.org:10.1056/NEJMoa012350>

65	Skyler, J. S. *et al.* Effects of oral insulin in relatives of patients with type 1 diabetes: The Diabetes Prevention Trial--Type 1. *Diabetes Care* **28**, 1068-1076 (2005). <https://doi.org:10.2337/diacare.28.5.1068>

66	Vehik, K. *et al.* Long-term outcome of individuals treated with oral insulin: diabetes prevention trial-type 1 (DPT-1) oral insulin trial. *Diabetes Care* **34**, 1585-1590 (2011). <https://doi.org:10.2337/dc11-0523>

67	Krischer, J. P., Schatz, D. A., Bundy, B., Skyler, J. S. & Greenbaum, C. J. Effect of Oral Insulin on Prevention of Diabetes in Relatives of Patients With Type 1 Diabetes: A Randomized Clinical Trial. *Jama* **318**, 1891-1902 (2017). <https://doi.org:10.1001/jama.2017.17070>

68	Elding Larsson, H., Lundgren, M., Jonsdottir, B., Cuthbertson, D. & Krischer, J. Safety and efficacy of autoantigen-specific therapy with 2 doses of alum-formulated glutamate decarboxylase in children with multiple islet autoantibodies and risk for type 1 diabetes: A randomized clinical trial. *Pediatr Diabetes* **19**, 410-419 (2018). <https://doi.org:10.1111/pedi.12611>

69	Herold, K. C. *et al.* An Anti-CD3 Antibody, Teplizumab, in Relatives at Risk for Type 1 Diabetes. *New England Journal of Medicine* **381**, 603-613 (2019). <https://doi.org:10.1056/nejmoa1902226>

70	Butty, V., Campbell, C., Mathis, D. & Benoist, C. Impact of diabetes susceptibility loci on progression from pre-diabetes to diabetes in at-risk individuals of the diabetes prevention trial-type 1 (DPT-1). *Diabetes* **57**, 2348-2359 (2008). <https://doi.org:10.2337/db07-1736>

71	Sosenko, J. M. *et al.* Slowed Metabolic Decline After 1 Year of Oral Insulin Treatment Among Individuals at High Risk for Type 1 Diabetes in the Diabetes Prevention Trial-Type 1 (DPT-1) and TrialNet Oral Insulin Prevention Trials. *Diabetes* **69**, 1827-1832 (2020). <https://doi.org:10.2337/db20-0166>

72	Hagopian, W. *et al.* Teplizumab preserves C-peptide in recent-onset type 1 diabetes: two-year results from the randomized, placebo-controlled Protege trial. *Diabetes* **62**, 3901-3908 (2013). <https://doi.org:10.2337/db13-0236>
